# Characterization of Age and Polarity at Onset in Bipolar Disorder

**DOI:** 10.1101/2021.04.16.21251163

**Authors:** Janos L. Kalman, Loes M. Olde Loohuis, Annabel Vreeker, Andrew McQuillin, Eli A. Stahl, Douglas Ruderfer, Maria Grigoroiu-Serbanescu, Georgia Panagiotaropoulou, Stephan Ripke, Tim B Bigdeli, Frederike Stein, Tina Meller, Susanne Meinert, Helena Pelin, Fabian Streit, Sergi Papiol, Mark J Adams, Rolf Adolfsson, Kristina Adorjan, Ingrid Agartz, Sofie R. Aminoff, Heike Anderson-Schmidt, Ole A. Andreassen, Raffaella Ardau, Jean-Michel Aubry, Ceylan Balaban, Nicholas Bass, Bernhard T Baune, Frank Bellivier, Antonio Benabarre, Susanne Bengesser, Wade H Berrettini, Marco P. Boks, Evelyn J Bromet, Katharina Brosch, Monika Budde, William Byerley, Pablo Cervantes, Catina Chillotti, Sven Cichon, Scott R Clark, Ashley L. Comes, Aiden Corvin, William Coryell, Nick Craddock, David W. Craig, Paul E. Croarkin, Cristiana Cruceanu, Piotr M. Czerski, Nina Dalkner, Udo Dannlowski, Franziska Degenhardt, J. Raymond DePaulo, Srdjan Djurovic, Howard J. Edenberg, Mariam Al Eissa, Torbjørn Elvsåshagen, Bruno Etain, Ayman H Fanous, Frederike Fellendorf, Alessia Fiorentino, Andreas J Forstner, Mark A. Frye, Janice M. Fullerton, Katrin Gade, Julie Garnham, Kirov George, Elliot Gershon, Michael Gill, Fernando S. Goes, Katherine Gordon-Smith, Paul Grof, Jose Guzman-Parra, Tim Hahn, Maria Hake, Roland Hasler, Urs Heilbronner, Stephane Jamain, Esther Jimenez, Ian Jones, Lisa Jones, Lina Jonsson, Rene S Kahn, John R. Kelsoe, James L. Kennedy, Tilo Kircher, Sarah Kittel-Schneider, Farah Klöhn-Saghatolislam, James A Knowles, Thorsten Manfred Kranz, Trine Vik Lagerberg, Mikael Landen, William B Lawson, Marion Leboyer, Qingqin S Li, Mario Maj, Dolores Malaspina, Mirko Manchia, Fermin Mayoral, Susan L McElroy, Melvin G McInnis, Andrew M McIntosh, Helena Medeiros, Ingrid Melle, Vihra Milanova, Philip B. Mitchell, Palmiero Monteleone, Alessio Maria Monteleone, Markus M Nöthen, Tomas Novak, John J Nurnberger, Niamh O’Brien, Kevin S. O’Connell, Claire O’Donovan, Michael C O’Donovan, Nils Opel, Abigail Ortiz, Michael J Owen, Erik Pålsson, Carlos Pato, Michele T Pato, Joanna Pawlak, Julia-Katharina Pfarr, Claudia Pisanu, James B. Potash, Mark H Rapaport, Daniela Reich-Erkelenz, Andreas Reif, Eva Reininghaus, Jonathan Repple, Helène Richard-Lepouriel, Marcella Rietschel, Kai Ringwald, Gloria Roberts, Guy Rouleau, Sabrina Schaupp, William A Scheftner, Simon Schmitt, Peter R. Schofield, K Oliver Schubert, Eva C. Schulte, Barbara Schweizer, Fanny Senner, Giovanni Severino, Sally Sharp, Claire Slaney, Olav B. Smeland, Janet L Sobell, Alessio Squassina, Pavla Stopkova, John Strauss, Alfonso Tortorella, Gustavo Turecki, Joanna Twarowska-Hauser, Marin Veldic, Eduard Vieta, John B. Vincent, Wei Xu, Clement C. Zai, Peter P. Zandi, Maria Del Zompo, Psychiatric Genomics Consortium (PGC) BD Working Group, International Consortium on Lithium Genetics (ConLiGen), Colombia-US Cross Disorder Collaboration in Psychiatric Genetics, Arianna Di Florio, Jordan W. Smoller, Joanna M. Biernacka, Francis J. McMahon, Martin Alda, Bertram Muller-Myhsok, Nikolaos Koutsouleris, Peter Falkai, Nelson B. Freimer, Till F.M. Andlauer, Thomas G Schulze, Roel A. Ophoff

## Abstract

**Background:** Studying the phenotypic and genetic characteristics of age and polarity at onset (AAO, PAO) in bipolar disorder (BD) can provide new insights into disease pathology and facilitate the development of screening tools.

**Aims:** To examine the genetic architecture of AAO and PAO and their association with BD disease characteristics.

**Methods:** Genome-wide association studies (GWASs) and polygenic score (PGS) analyses of AAO (N=12977) and PAO (N=6773) were conducted in BD patients of 34 cohorts and a replication sample (N=2237). The association of onset with disease characteristics was investigated in two of these cohorts.

**Results:** Earlier AAO was associated with an increased risk of psychotic symptoms, suicidality, and fewer episodes. A depressive onset correlated with lifetime suicidality and a manic onset with delusions and manic episodes. Systematic differences in AAO between cohorts and continents of origin were observed. This was also reflected in SNV-based heritability estimates, with higher heritabilities for stricter onset definitions. Increased polygenic scores for autism spectrum disorder (β=-0.34 years, SE=0.08), major depression (β=-0.34 years, SE=0.08), schizophrenia (β=-0.39 years, SE=0.08), and educational attainment (β=-0.31 years, SE=0.08) were associated with an earlier AAO. The AAO GWAS identified one significant locus, but this finding did not replicate. Neither GWAS nor PGS analyses yielded significant associations with PAO.

**Conclusions:** AAO and PAO are associated with indicators of BD severity. Individuals with an earlier onset show an increased polygenic liability for a broad spectrum of psychiatric traits. Systematic differences in AAO across cohorts, continents, and phenotype definitions introduce significant heterogeneity, affecting analyses.

**RELEVANCE STATEMENT:** In the largest study to systematically characterize age at onset (N=12977) and polarity at onset (N=6773) in bipolar disorder, we describe an association between illness onset characteristics and indicators of severity, confirming their clinical relevance. Our study shows that that early illness onset is associated with genetic liability for a broad range of psychiatric disorders. However, we also highlight systematic differences in age at onset across cohorts, continents, and phenotype definitions. This heterogeneity results in reduced heritability and affects genetic analyses, underscoring the need for the development of standardized phenotype definitions.

## 1. Introduction

Bipolar disorder (BD) is highly heritable and affects approximately 1% of the population (1). It has a recurrent or chronic course and is associated with psychosocial impairment and reduced functioning, and it is a leading cause of global disease burden (2,3). Individuals usually experience their first (hypo)manic or depressive episode of BD in adolescence or early adulthood, but often they are not diagnosed until five to ten years later (4), especially in case of earlier age at onset or a depressive index episode (5). Early illness onset is associated with a more severe disease course and greater impairment across a wide range of mental and physical disorders and is a useful prognostic marker (6–9). However, pathophysiological processes leading to a disorder are thought to begin long before the first symptoms appear (10,11). Investigating the factors contributing to age and polarity (i.e., either a (hypo)manic or depressive episode) at onset could thus improve our understanding of disease pathophysiology and facilitate development of personalized screening and preventive measures. Accordingly, age and polarity at BD onset are considered as suitable phenotypes for genetic analyses. Genome-wide association studies (GWASs) have improved our understanding of the genetic architecture of susceptibility to BD; however, the genetic determinants of age and polarity at onset remain largely unknown. Evidence suggests that patients with an early age at onset carry a stronger genetic loading for BD risk (12). For example, an earlier parental age at onset increases familial risk for BD and is one of the strongest predictors of 5-year illness onset in affected offspring (12–14). Thus far, GWASs for age at BD onset have been underpowered (15–17), and a study on 8610 cases found no significant evidence for a heritable component contributing to onset age (16). Previous research suggested that a higher genetic risk burden for schizophrenia (SZ) may be associated with earlier age at onset of BD (16), but this finding was not replicated (18–20). Moreover, a recent study did not find an association of BD PGS with age at onset (21). Polarity of onset was shown to cluster in families (22), but the genetic architecture of polarity at onset has not yet been investigated. To fill these knowledge gaps, we performed comprehensive analyses of age and polarity at BD onset in the largest sample studied to date by (i) examining phenotype definitions and associations, (ii) conducting systematic GWASs, and (iii) investigating whether genetic load for neuropsychiatric disorders and traits contributes to age and polarity at BD onset.

## 2. Methods

### 2.1. Sample for the discovery GWAS

Participants with a BD diagnosis and age at onset information were selected from independent datasets, including those previously submitted to the Psychiatric Genomics Consortium (PGC) BD Working Group (18,23) and the International Consortium on Lithium Genetics (ConLiGen) (24). These consortia aggregate data from many cohorts worldwide. Our analyses comprised 34 cohorts with 12977 BD European ancestry cases from Europe, North America, and Australia. For a description of sample ascertainment, see the Supplementary Material. The authors assert that all procedures contributing to this work comply with the ethical standards of the relevant national and institutional committees on human experimentation and with the Helsinki Declaration of 1975, as revised in 2008. All procedures involving human subjects/patients were approved by the local ethics committees, and written informed consent was obtained from all subjects/patients.

#### 2.1.1. Definition of age at onset

The definition of age at BD onset differed by cohort. To enhance cross-cohort comparability, we grouped the definitions into four broad categories (*Supplementary Table S1*): (1) *Diagnostic interview*: age at which the patient first experienced a (hypo)manic, mixed, or major depressive episode according to a standardized diagnostic interview; (2) *Impairment/help-seeking*: age at which symptoms began to cause subjective distress or impaired functioning or at which the patient first sought psychiatric treatment; (3) *Pharmacotherapy*: age at first administration of medication; (4) *Mixed*: a combination of the above-mentioned definitions. Across definitions, participants younger than eight years at onset were excluded (n=279) because of the uncertainty about the reliability of retrospective recall of early childhood onset.

The distribution of age at onset was highly skewed and differed greatly across the cohorts *(Table 1 and Fig. 1)*. Therefore, we transformed age at onset in each cohort by rank-based inverse normal transformation and used this normalized variable as the primary dependent variable in all genetic analyses. To facilitate interpretability of effect sizes, we also report results of the corresponding untransformed age at onset.

**Table 1.** Sample characteristics of datasets used in genetic analyses

| GWAS stage | Dataset | N | Continent | Diagnosis, % BD-I | Sex, % male | AAO, median (MAD<sup>+</sup>, range) | Definition of AAO | PAO*, N (%) |
| --- | --- | --- | --- | --- | --- | --- | --- | --- |
| Discovery | wtccc | 1452 | Europe | 89.53% | 36.85% | 24 (8.9, 9-63) | Impairment / help-seeking |  |
|  | tgco2 | 865 | North America | 100% | 33.64% | 17 (5.93, 8-46) | Diagnostic interview | PAO-M: 316 (38.92%);<br>PAO-D: 496 (61.08%) |
|  | gain | 797 | North America | 100% | 48.06% | 18 (5.93, 8-45) | Diagnostic interview | PAO-M: 135 (18.57%);<br>PAO-D: 440 (60.52%) |
|  | stp1 | 718 | North America | 100% | 44.01% | 16 (5.93, 8-41) | Diagnostic interview | PAO-M: 137 (19.08%);<br>PAO-D: 420 (58.5%) |
|  | gsk1 | 715 | North America | 89.51% | 36.36% | 19 (7.51, 8-52) | Diagnostic interview | PAO-M: 102 (14.61%);<br>PAO-D: 395 (56.59%) |
|  | usc2 | 681 | North America | 96.18% | 47.58% | 18 (7.41, 8-48) | Impairment / help-seeking |  |
|  | bonn | 638 | Europe | 99.84% | 47.34% | 25 (8.9, 9-64) | Impairment / help-seeking |  |
|  | ucl2 | 604 | Europe | 100% | 44.37% | 30 (11.86, 9-60) | Pharmacotherapy | PAO-M: 47 (9.96%);<br>PAO-D: 209 (44.28%) |
|  | bmg3 | 455 | Europe | 57.14% | 40.66% | 24 (10.38, 10-62) | Impairment / help-seeking | PAO-M: 43 (16.35%);<br>PAO-D: 159 (60.46%) |
|  | m&m's | 449 | Europe | 74.83% | 52.12% | 23 (10.38, 8-65) | Mixed | PAO-M: 73 (17.14%);<br>PAO-D: 238 (55.87%) |
|  | uclo | 439 | Europe | 100% | 39.86% | 22 (7.41, 8-51) | Impairment / help-seeking | PAO-M: 54 (14.25%);<br>PAO-D: 197 (51.98%) |
|  | fran | 411 | Europe | 77.62% | 41.36% | 22 (7.41, 10-58) | Diagnostic interview |  |
|  | euoR | 410 | Europe | 75.85% | 44.15% | 22 (9.64, 11-59) | Mixed |  |
|  | hal2 | 355 | North America | 71.55% | 42.54% | 23 (8.9, 8-56) | Diagnostic interview | PAO-M: 102 (29.65%);<br>PAO-D: 213 (61.92%) |
|  | ume4 | 354 | Europe | 69.21% | 37.85% | 20 (8.9, 8-63) | Diagnostic interview | PAO-M: 54 (14.25%);<br>PAO-D: 197 (51.98%) |
|  | swa2 | 344 | Europe | 81.10% | 41.86% | 23 (10.38, 10-70) | Impairment / help-seeking |  |
|  | bmpo | 319 | Europe | 78.06% | 39.18% | 28 (11.86, 10-63) | Impairment / help-seeking | PAO-M: 41 (16.33%);<br>PAO-D: 150 (59.76%) |
|  | **top7** | 301 | Europe | 62.79% | 41.53% | 19 (7.41, 8-49) | Diagnostic interview |  |
|  | **may1** | 257 | North America | 100% | 45.14% | 20 (8.9, 8-62) | Diagnostic interview | PAO-M: 34 (13.23%);<br>PAO-D: 142 (55.25%) |
|  | **bmsp** | 248 | Europe | 94.76% | 45.56% | 22 (7.41, 9-57) | Impairment / help-seeking | PAO-M: 24 (10.04%);<br>PAO-D: 93 (38.91%) |
|  | **bmau** | 245 | Australia | 79.18% | 40.82% | 19 (7.41, 8-55) | Diagnostic interview | PAO-M: 46 (20.18%);<br>PAO-D: 125 (54.82%) |
|  | **edi1** | 244 | Europe | 99.18% | 42.62% | 20 (5.93, 13-50) | Diagnostic interview |  |
|  | **rom3** | 226 | Europe | 100% | 41.15% | 25 (10.38, 12-59) | Diagnostic interview | PAO-M: 91 (40.27%);<br>PAO-D: 134 (59.29%) |
|  | **butr** | 204 | Europe | 100% | 40.2% | 22 (5.19, 13-44) | Impairment / help-seeking |  |
|  | **euol** | 191 | Europe | 74.87% | 31.41% | 24 (8.9, 13-67) | Diagnostic interview | PAO-M: 48 (27.43%);<br>PAO-D: 98 (56%) |
|  | **ageu** | 178 | Europe | 90.45% | 39.33% | 21 (7.41, 8-51) | Impairment / help-seeking |  |
|  | **mich** | 169 | North America | 100% | 31.36% | 18 (5.93, 8-45) | Diagnostic interview | PAO-M: 42 (24.85%);<br>PAO-D: 84 (49.7%) |
|  | **naom** | 159 | North America | 84.91% | 44.65% | 18 (7.41, 8-66) | Mixed | PAO-M: 30 (28.85%);<br>PAO-D: 51 (49.04%) |
|  | **bmg2** | 152 | Europe | 59.87% | 35.53% | 27 (10.38, 13-63) | Impairment / help-seeking |  |
|  | **top8** | 111 | Europe | 55.86% | 37.84% | 18 (7.41, 8-49) | Diagnostic interview |  |
|  | **h66x** | 92 | Europe | 82.61% | 36.96% | 30 (10.38, 9-55) | Mixed |  |
|  | **auom** | 85 | Australia | 88.24% | 45.88% | 25 (10.38, 8-64) | Diagnostic interview |  |
|  | **euo2** | 58 | Europe | 65.52% | 56.9% | 26 (8.9, 18-57) | Diagnostic interview |  |
|  | **dub1** | 51 | Europe | 100% | 54.9% | 21 (5.93, 12-45) | Diagnostic interview |  |
|  | **Summary** | **12977** |  | **88.27%** | **41.57%** | **21 (8.9, 8-70)** |  | **PAO-M: 1435 (21.19%);**<br>**PAO-D: 3885 (57.36%)** |
| **Replication** | **ukwa1** | 1156 | Europe | 75.17% | 38.15% | 23 (8.9, 8-74) | Impairment / help-seeking |  |
|  | **dutch** | 468 | Europe | 100% | 42.31% | 28 (10.38, 11-63) | Pharmacotherapy |  |
|  | **jst5** | 186 | North America | 100% | 53.23% | 16 (7.41, 8-51) | Unknown |  |
|  | **colo** | 176 | South America | 90.34% | 31.82% | 20 (11.86, 8-52) | Diagnostic interview |  |
|  | **bmrom** | 126 | Europe | 100% | 42.86% | 24 (8.9, 12-56) | Diagnostic criteria |  |
|  | **bdtrs** | 125 | Europe | 64% | 45.6% | 28 (13.34, 8-65) | Impairment / help-seeking |  |
|  | **Summary** | **2237** |  | **84.40%** | **40.46%** | **24 (10.38, 8-74)** |  |  |
| **All data** |  | **15214** |  | **86.26%** | **41.41%** | **22 (8.9, 8-74)** |  |  |

*Abbreviations: GWAS, genome-wide association study; BD-I, bipolar disorder type I., AAO, age at onset; MAD, mean absolute* *deviation, PAO, polarity at onset;*
*\*We calculated the mean absolute deviation using 1.4826 as constant.*
*\*We defined three categories of polarity at onset: PAO-M, mania/hypomania before depression; PAO-D, depression before* *mania/hypomania; and PAO-X, mixed. PAO was not available for all patients. The table presents the PAO-M and PAO-D subgroups* *and their percentage within the individual cohorts.*

**Figure 1:**
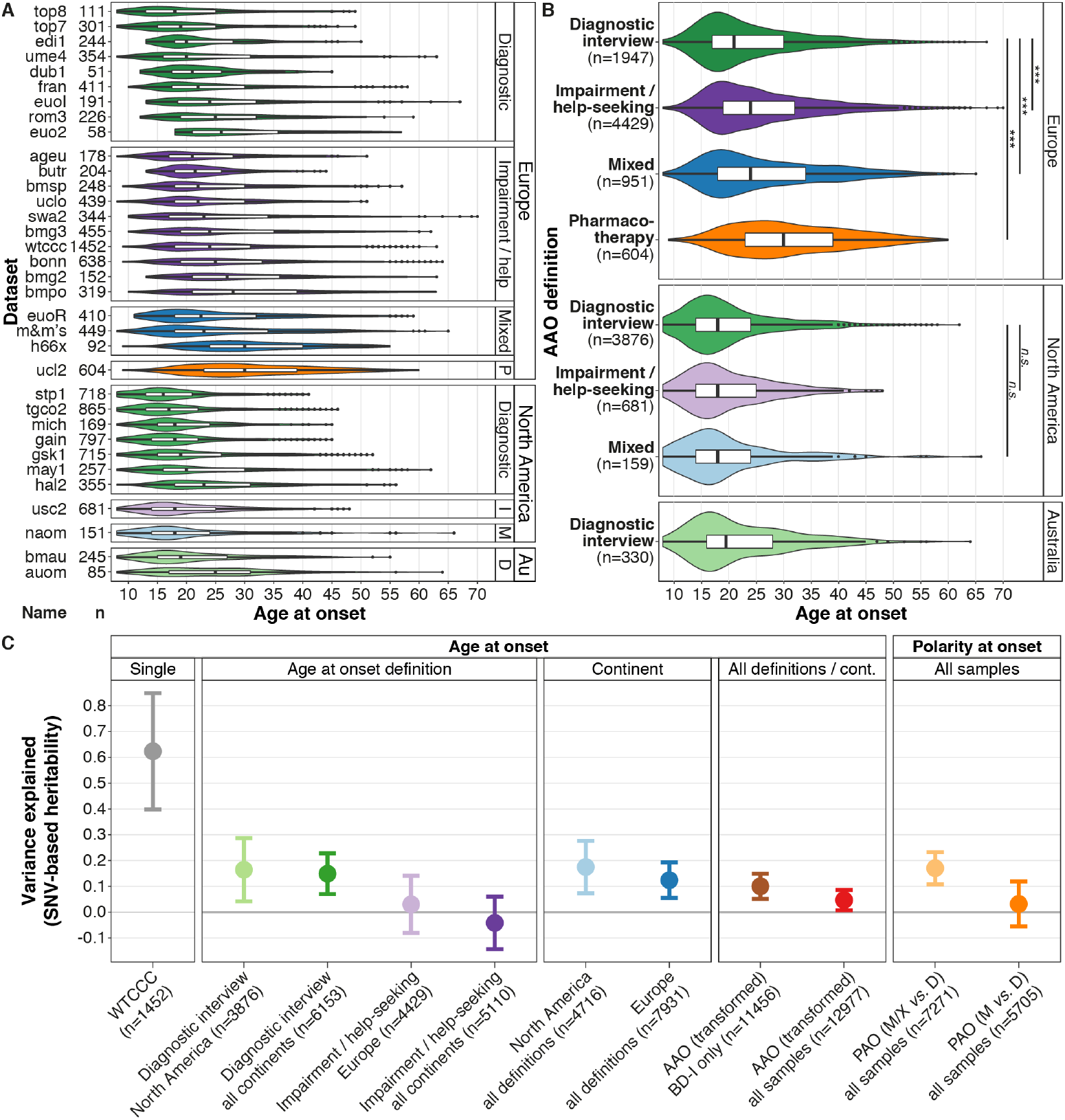
Differences between phenotype definitions and continents across the 34 datasets used for genetic analyses **A:** The various datasets used four different definitions for age at onset: diagnostic interview, impairment/help-seeking, pharmacotherapy, and mixed. **B:** The untransformed age at onset differed significantly, depending on the phenotype definition used. Abbreviations: P, pharmacotherapy; I, impairment/help-seeking; M, mixed; D, diagnostic interview; *n*.*s*., *P*>0.05; *** *P*<0.001. **C:** Estimation of the variance in different phenotype definitions explained by genotyped single-nucleotide variants (*h*^*2*^_*SNV*_). For the cohort *wtccc*, we directly estimated *h*^*2*^_*SNV*_ from genotype data in GCTA GREML; we estimated all other heritabilities from GWAS summary statistics in LDSC. The plot shows *h*^*2*^_*SNV*_ estimates and SEs. Abbreviations: SNV, single-nucleotide variant; cont, continent; AAO, age at onset; BD-I, BD type I; PAO, polarity at onset; PAO-M, mania/hypomania before depression; PAO-D, depression before mania/hypomania; PAO-X, mixed.

#### 2.1.2. Definition of polarity at onset

For each cohort, polarity at onset was defined by comparing the age at the first (hypo)manic and first depressive episode or using the polarity variable provided by the cohort. Specifically, patients were divided into three subgroups: (1) (hypo)mania before depression (PAO-M), (2) depression before (hypo)mania (PAO-D), and (3) mixed (PAO-X). The third category included patients with mixed episodes and those with a first (hypo)manic and depressive episode within the same year *(Table 1)*. In the primary analysis, we combined (hypo)mania and mixed onset cases and assigned this as the reference category. In secondary analyses, we excluded the mixed cases.

### 2.2. Disease characteristics

We performed phenotypic analyses of disease onset in BD type I patients from three cohorts: the Dutch Bipolar cohort (N=1313) (25) and the German PsyCourse (26) and FOR2107 (27) cohorts, which were analyzed jointly (N=346). We analyzed the following disease characteristics, which were previously reported as being associated with disease onset and were assessed in a similar way across cohorts (28): lifetime delusions, lifetime hallucinations, history of suicide attempt, suicidal ideation, current smoking, educational attainment, living together with a partner, and frequency of manic and depressive episodes per year. For more detailed information, see the *Supplementary Material* and *Supplementary Table S9*.

### 2.3. Genetic analyses

We analyzed age at onset in 34 cohorts comprising 12977 BD patients. We performed a GWAS on these cohorts and attempted to replicate any findings in a GWAS on six additional cohorts with 2237 BD patients. For details on the datasets, including phenotype definitions and distributions, see *Table 1, Fig. 1*, and *Supplementary Table S1*.

#### 2.3.1. Genotype data quality control and imputation

Cohorts were genotyped according to local protocols. Individual genotype data of all discovery-stage cohorts were processed with the PGC Rapid Imputation and Computational Pipeline for GWAS (RICOPILI) (29) with the default parameters for standardized quality control (QC), imputation, and analysis. Before imputation, filters for the removal of variants included non-autosomal chromosomes, missingness ≥0.02, and a Hardy-Weinberg equilibrium test P <1×10^−10^. Individuals were removed if they showed a genotyping rate ≤0.98, absolute deviation in autosomal heterozygosity of Fhet ≥0.2, or a deviation >4 SDs from the mean in any of the first eight multidimensional scaling (MDS) ancestry components within each cohort. From genetic duplicates (assessed with the genetic identity-by-state matrix) and relatives (pi-hat >0.2) across all samples, only the case with more complete phenotypic information on AAO and PAO, sex, and diagnosis was retained. Imputation was performed by IMPUTE2 with the Haplotype Reference Consortium reference panel (30).

#### 2.3.2. Genome-wide association studies

We conducted separate GWASs for all cohorts containing 40 or more individuals in RICOPILI/PLINK (29,31), with sex, BD subtype, and the first eight ancestry components as covariates. Sample sizes are provided in *Supplementary Tables S2, S7*. The results showed no inflation of test statistics *(Supplementary Table S2)*. We combined the GWASs in METAL by fixed-effects meta-analysis (32). The primary analyses were transformed age at onset (analyzed by linear regression), and polarity at onset (analyzed by logistic regression). Secondary analyses included GWASs stratified by age at onset definition and continent of origin.

For the meta-analysis summary statistics, we applied the following variant-level post-QC parameters: imputation INFO score□≥0.9, MAF□≥0.05, and successfully imputed/genotyped in□more than□half of the cohorts.

We estimated power to replicate our original GWAS finding using the *pwr* package in R. Given the parameters from the locus identified through our discovery GWAS (beta =0.075, allele frequency =0.32) and a standardized phenotype, we had 76% power to detect the effect in our sample size of 2237 at an alpha level of 0.1. For comparison, we had 57% power to detect the effect in our discovery sample.

#### 2.3.3. Polygenic Scoring

We calculated polygenic scores (PGS) based on prior GWAS of Attention-Deficit/Hyperactivity Disorder (ADHD) (33), Autism Spectrum Disorder (ASD) (34), BD (18), Educational Attainment (EA, measured as “years in education”) (35), Major Depression (MD) (36), and schizophrenia (37) *(Supplementary Table S3)*. PGS weights were estimated with PRS-CS (38), with six scores per genome-wide association study (GWAS; with φ = 1×10^−1^, 1×10^−2^, 1×10^−3^, 1×10^−4^, 1×10^−5^, and 1×10^−6^). We tested the associations of the PGS with the AAO and PAO by linear and logistic regressions, respectively, with the same covariates as in the GWAS analyses. The significance threshold was Bonferroni-corrected for 96 tests (α =□0.05/[6 φ thresholds × 8 traits × 2 phenotypes] =5.2×10^−4^).

#### 2.3.3. Heritability analyses

We assessed the variance in age and polarity at onset explained by genotyped variants (single-nucleotide variant [SNV]-based heritability, *h*^*2*^ _*SNV*_). For individual cohorts with more than 1000 samples, we estimated *h*^*2*^ _*SNV*_ with GCTA GREML (39,40). Here, we validated the robustness of the *h*^*2*^ _*SNV*_ estimate with the mean of 1000× resampling of 95% of the sample. For meta-analysis summary statistics with sample sizes >3000, we estimated *h*^*2*^ _*SNV*_ by linkage disequilibrium (LD) score regression (41). The 95% CIs were constrained to a minimum of 0 and a maximum of 1.

## 3. Results

### 3.1. Heterogeneity of age and polarity at onset across cohorts

Among the four definitions of age at onset across the 34 cohorts, impairment/help-seeking was the most common in Europe and diagnostic interview the most common in North America *(Table 1, Fig. 1A-B)*. Across cohorts, the median age at onset was 21 years (range of medians: 16-30 years; *Fig. 1A-B*). The median untransformed age at onset differed significantly between BD subtypes (type I, 21 years; type II, 22 years; Kruskal-Wallis test *P*=1.8×10^−4^; *Supplementary Table S6*), age at onset definitions (diagnostic interview, 19 years; impairment/help-seeking, 23 years; pharmacotherapy, 30 years; mixed, 22 years; *P*=2.96×10^−191^), and continent of origin (Europe, 24 years; North America, 18 years; and Australia, 19.5 years; *P*=2.0×10^− 263^). These differences across continents remained significant when including onset definitions and BD subtype in a multivariable regression model, indicating that they are not entirely driven by differences between diagnostic instruments *(Supplementary Table S6)*.

The majority of patients had a depression-first polarity at onset in both Europe and North America (57% and 59%, respectively; *P*=0.17 test of proportion). Depression-first cases were less frequent in the impairment/help-seeking than in the diagnostic interview category (55% and 60%, respectively; *P*=4.5×10^−4^, *Supplementary Fig. S1*).

### 3.2. Analyses of disease characteristics

In a meta-analysis of the Dutch and German samples, earlier age at onset was significantly associated with a higher probability of lifetime delusions, hallucinations, suicide attempts, suicidal ideation, lower educational attainment, and not living together *(Table 2, Supplementary Tables S4 and S5)*. Later age at onset was positively significantly correlated with a higher number of manic and depressive episodes per year (see secondary analyses in the *Supplementary Material*). Moreover, a (hypo)manic onset was significantly associated with a greater likelihood of delusions and more manic episodes per year, whereas a depressive onset was associated with a higher probability of suicidal ideation and lifetime suicide attempts.

**Table 2.** The association of age and polarity at onset with disease characteristics in two European BD cohorts

| Disease characteristic | AAO |  |  |  |  | PAO |  |  |  |  |
| --- | --- | --- | --- | --- | --- | --- | --- | --- | --- | --- |
|  | N | Odds ratio | 95% CI | Unadjusted P value | Adjusted P value* | N | Odds ratio | 95% CI | Unadjusted P value | Adjusted P value |
| Delusions | 1612 | 0.71 | 0.64-0.79 | $1.61 \times 10^{-9}$ | **$1.45 \times 10^{-8}$** | 1298 | 0.62 | 0.49-0.79 | $1.04 \times 10^{-4}$ | **$6.24 \times 10^{-4}$** |
| Hallucinations | 1594 | 0.83 | 0.74-0.92 | $3.5 \times 10^{-4}$ | **$1.40 \times 10^{-3}$** | 1290 | 0.93 | 0.74-1.17 | $5.22 \times 10^{-1}$ | $1.00 \times 10^0$ |
| Current smoking | 1594 | 0.98 | 0.89-1.09 | $7.50 \times 10^{-1}$ | $7.50 \times 10^{-1}$ | 1282 | 1.12 | 0.89-1.41 | $3.39 \times 10^{-1}$ | $1.00 \times 10^0$ |
| Suicidal ideation | 1518 | 0.79 | 0.71-0.88 | $2.31 \times 10^{-5}$ | **$1.62 \times 10^{-4}$** | 1280 | 1.68 | 1.32-2.13 | $2.11 \times 10^{-5}$ | **$1.48 \times 10^{-4}$** |
| Suicide attempt | 1537 | 0.78 | 0.69-0.88 | $2.73 \times 10^{-5}$ | **$1.64 \times 10^{-4}$** | 1262 | 1.58 | 1.24-2.02 | $2.67 \times 10^{-4}$ | **$1.34 \times 10^{-3}$** |
| Educational attainment | 1636 | 1.17 | 1.06-1.29 | $2.77 \times 10^{-3}$ | **$8.31 \times 10^{-3}$** | 1319 | 1.06 | 0.85-1.33 | $5.93 \times 10^{-1}$ | $1.00 \times 10^0$ |
| Living together | 1357 | 1.28 | 1.15-1.44 | $1.01 \times 10^{-5}$ | **$8.08 \times 10^{-5}$** | - | - | - | - | - |
|  |  | Estimate** | SE | Unadjusted P value | Adjusted P value* |  | Estimate | SE | Unadjusted P value | Adjusted P value |
| Number of manic episodes per illness year | 1436 | 0.11 | 0.03 | $7.08 \times 10^{-5}$ | **$3.54 \times 10^{-4}$** | 1156 | -0.42 | 0.06 | $4.68 \times 10^{-13}$ | **$3.74 \times 10^{-12}$** |
| Number of depressive episodes per illness year | 1231 | 0.07 | 0.03 | $1.93 \times 10^{-2}$ | **$3.86 \times 10^{-2}$** | 1051 | 0.12 | 0.06 | $4.63 \times 10^{-2}$ | $1.85 \times 10^{-1}$ |

Abbreviations: AAO, age at onset; PAO, polarity at onset
N: total number of participants from the Dutch and German cohorts
The number of manic/depressive episodes was divided by (years of illness)+1. For secondary analyses of the number of episodes not corrected for the years of illness, see the Supplementary Material.
\* After Bonferroni-Holm correction. Significant adjusted P values are indicated in bold.
\*\* Unstandardized beta coefficient

### 3.3. Genome-wide association studies

In our GWAS meta-analysis of age at onset using all 34 cohorts, one locus reached genome-wide significance (rs1610275 on chromosome 16; minor allele G frequency=0.319, β=0.075 SDs, SE=0.014, *P*=3.39×10^−8^, *Fig. 2A, Supplementary Table S7, Supplementary Fig. S2*). This SNV mapped to an intron of the brain-expressed gene *FTO* (alpha-ketoglutarate dependent dioxygenase, *Fig. 2B*). However, this association was not replicated in an independent sample of six cohorts *(Supplementary Table S7, Supplementary Fig. S2)*. In this replication sample (N=2237), we had 76% power to replicate this SNV at *P*=0.1.

**Figure 2:**
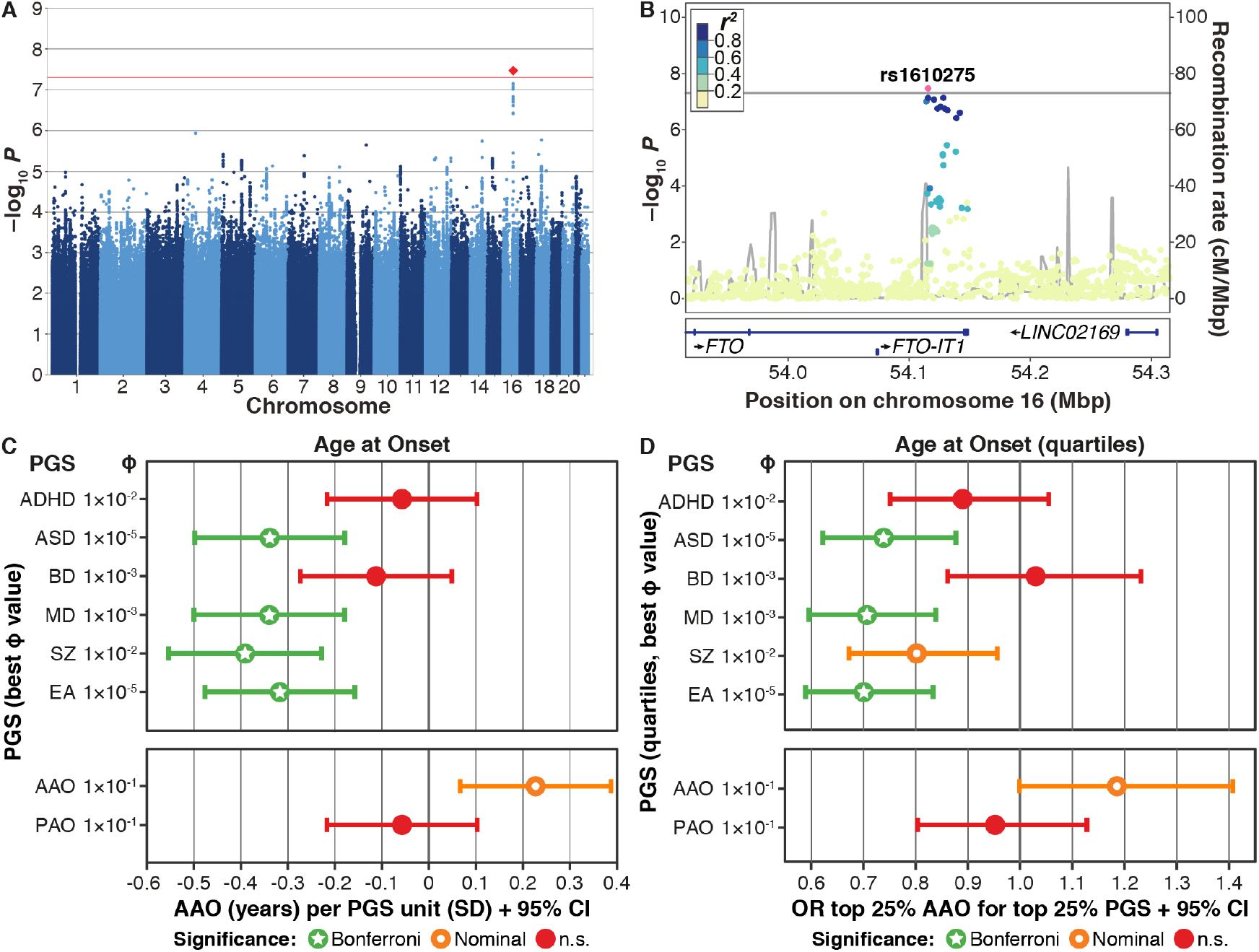
Results from the genome-wide association study (GWAS) and polygenic score (PGS) analyses of bipolar disorder (BD) age at onset (AAO) **A:** Manhattan plot of the discovery-stage GWAS. **B:** Locus-specific Manhattan plot of the top-associated variant. Mbp, mega base pairs; cM, centi Morgan. **C-D:** Results from analyses of PGS. For detailed results, see Supplementary Table S8. Significance levels: n.s., *P*>0.05; Nominal, *P*<0.05; Bonferroni, below the Bonferroni-corrected significance threshold corrected for 96 tests (*P*<5.2×10^−4^). Abbreviations: ADHD, attention deficit/hyperactivity disorder; ASD, autism spectrum disorder; MD, major depression; SZ, schizophrenia; EA, educational attainment. **C:** For interpretability, the plot shows the untransformed AAO. Significance levels are based on the analyses of the AAO after rank-based inverse normal transformation (which was performed because the distribution of AAO was highly skewed and differed greatly across the study cohorts). **D:** Associations of the top vs bottom AAO quartiles with the top vs bottom PGS quartiles. A higher odds ratio (OR) indicates an association with higher AAO.

In the GWAS of polarity at onset, no variant was genome-wide significant in either primary (PAO-M/-X *vs* PAO-D) or secondary (PAO-M *vs* PAO-D) analyses *(Supplementary Fig. S3)*.

We calculated PGSs for age and polarity at onset using leave-one-out summary statistics from these GWASs. The age-at-onset PGS was nominally significantly associated with age at onset (β=0.23 years, SE=0.08, *P*=0.0087, φ=0.1, *Fig. 2C-D*) for five of six tested φ parameters but did not withstand correction for multiple testing *(Supplementary Table S8)*. The polarity-at-onset PGS was not associated with the polarity at onset *(Supplementary Fig. S4)*.

### 3.4. SNV-based heritability of the investigated phenotypes

Using the GWAS summary statistics, we estimated the SNV-based heritability *h*^*2*^_*SNV*_ by LD score regression *(Fig. 1C)*. We estimated *h*^*2*^_*SNV*_ directly from genotype data in the only cohort large enough for this analysis, *wtccc h*^*2*^_*SNV*_ =0.63, *P*=0.0026) and validated the robustness of this estimate by resampling (*h*^*2*^_*SNV*_ =0.62, resampling 95% CI=0.15-1.00). The *h*^*2*^_*SNV*_ estimates decreased when cohorts, phenotype definitions, and continents were combined (e.g., “diagnostic interview” in North America: *h*^*2*^_*SNV*_=0.16 [95% CI=0-0.40], “impairment/help seeking” in Europe: *h*^*2*^ =0.03 [95% CI=0-0.25], all combined *h*^*2*^_*SNV*_=0.05 [95% CI=0-0.12]). Because the sample sizes were smaller, we could not estimate the *h*^*2*^_*SNV*_ of impairment/help seeking in North America and diagnostic interview in Europe. For depressed *vs*. (hypo)manic and mixed polarity at onset, *h*^*2*^_*SNV*_ was 0.17 (95% CI=0.05-0.29) on the observed scale.

### 3.5. Associations of PGSs with age and polarity at onset

We analyzed whether PGSs for five psychiatric disorders and EA were associated with earlier age at onset *(Fig. 2C-D, Supplementary Table S8)*. After correcting for 96 tests, higher PGSs for ASD (β=-0.34 years per 1 SD increase in PGS, SE=0.08, *P*=9.85×10^−6^), MD (β=-0.34, SE=0.08, *P*=1.40×10^−6^), SZ (β=-0.39, SE=0.08, *P*=2.91×10^−6^), and EA (β=-0.31, SE=0.08, *P*=5.58×10^−5^) were significantly associated with an earlier BD age at onset. This was not the case for ADHD or BD PGS. No PGS was significantly associated with polarity at onset *(Supplementary Fig. S4, Supplementary Table S8)*.

## 4. Discussion

Our study represents the largest and most comprehensive characterization of disease onset in BD to date. It highlights the clinical relevance of age and polarity at onset in BD by demonstrating how these phenotypes are associated with several indicators of illness severity. We found that higher PGS for ASD, MD, SZ, and EA were associated with an earlier age at onset. Thereby we demonstrated that the BD age at onset has a polygenic etiology. Individuals with an earlier age at onset harbor increased genetic liability for a broad spectrum of psychiatric disorders and related traits. However, we show significant heterogeneity of age at onset (and, partly, polarity at onset) across cohorts, continents, and age-at-onset definitions and demonstrate the challenges for genetic analyses that occur in the presence of substantial phenotypic heterogeneity. First, heritability estimates differed for criteria used to define age at onset and decreased when combining multiple cohorts. Second, a single genome-wide significant variant, identified in the age-at-onset discovery GWAS, did not replicate in an independent dataset.

A striking finding of our study was the systematic difference in the age at onset distribution across cohorts, continents, and assessment strategies. The definition of age at onset correlated strongly with the continent, with diagnostic interview being mainly used in North America and impairment/help-seeking in Europe. Using multivariable regression, we showed that the continent-level differences cannot fully be explained by the age-at-onset assessment strategy and that both factors contribute significantly to the heterogeneity (Supplementary Table S6) (4). However, variations in the demographic structure of analyzed populations may have biased the assessed age at onset of BD, contributing to the observed differences (42). Although prior research has identified age at onset differences across continents (e.g., the incidence of early onset BD is higher in the US than in Europe (43,44)), this study is the first to systematically assess this heterogeneity across many cohorts with different ascertainment strategies and demonstrate how it may reduce statistical power in genetic analyses.

Interestingly, these systematic differences in age at onset phenotypes are reflected in heritability estimates: We observed the highest SNV-based heritability *h*^*2*^_*SNV*_when onset was established by diagnostic interview and the lowest when it was captured with more health system-specific and subjective measurements, such as item 4 of the Operational Criteria Checklist for Psychotic Illness (impairment/help-seeking). Moreover, *h*^*2*^_*SNV*_ estimates approached zero when all samples were combined in our primary analysis (h^2^_*SNV*_=0.05; 95% CI=0-0.12), underscoring the strong impact of phenotypic heterogeneity. Thus, we not only showed systematic heterogeneity in a clinically relevant psychiatric phenotype across cohorts but also provided direct evidence for how this heterogeneity can hamper genetic studies. Notably, a recent investigation demonstrated that the phenotyping method (e.g., diagnostic interview vs. self-report) significantly influenced heritability estimates, GWAS results, and PGS performance in analyses of MD susceptibility, with broader phenotype definitions resulting in lower heritability estimates (45). Nevertheless, the significant results in our PGS association analyses confirmed that a significant genetic component of age at onset exists.

For polarity at disease onset, the relative proportion of patients reporting a depressive index episode did not differ across continents but did differ across instruments, i.e., a (hypo)manic onset was more common if age at onset was based on impairment/help-seeking instead of diagnostic interview.

Although our discovery GWAS identified a genome-wide significant locus for age at onset, the lack of replication suggests that this finding may have been a false-positive. Of note, our replication power analysis did not account for the phenotypic and genetic heterogeneity across the cohorts and may thus have underestimated the necessary sample size. Moreover, the replication sample was more ethnically diverse than the discovery sample, reducing statistical power. While the effect of the age at onset PGS on the trait measure itself was substantial (0.23 years per 1 SD change in the PGS), the association was only nominally significant.

Possibly, the rank-based inverse-normal transformation of the age at onset affected the GWAS and heritability analyses. We conducted this transformation because, first, the original age at onset distribution was highly skewed and thus not suitable for linear regression and, second, the age at onset differed significantly between cohorts, which could have biased the meta-analysis. By transforming the data, however, only the rank and not the absolute differences in onset between patients was maintained, reducing interpretability of the phenotype and the genetic effects.

Higher PGSs for SZ, MD, ASD, and EA were significantly associated with a lower age at onset, confirming a genetic component for age at onset. This finding supports the hypothesis that earlier age of onset, in addition to potentially being driven by onset-specific modifier genes, is influenced by a transdiagnostic liability for psychiatric disorders. In contrast to several other disorders (e.g. multiple sclerosis and late-onset Alzheimer’s disease), where the strongest genetic risk factors for disease liability were also the most important genetic factors associated with earlier disease onset (8,46), we did not find a significant association between BD PGS and age at BD onset, confirming a previous study (21). Importantly, the sample sizes of both the SZ and MD GWASs were larger than those of the BD GWAS, improving the predictive power of these PGSs compared to the BD PGS.

Our finding of a significant relationship of higher EA PGS with an earlier age at onset may seem counterintuitive. However, several studies found a significant association between higher EA and BD (47,48). In addition, a recent study demonstrated a positive genetic correlation and causal relationship between high EA and BD (49). Our findings show that high EA is not only a risk factor for BD but is also associated with an earlier onset.

In our analyses of polarity at onset, the small sample size and dichotomous phenotype reduced statistical power. None of the tested PGSs were significantly associated with polarity at onset, and the GWAS did not identify genome-wide significant loci. However, we observed significant *h*^*2*^ estimates for PAO-M/-X *vs*. PAO-D, providing substantial evidence that genetic factors contribute to the polarity at BD onset.

The success of GWASs in the study of complex, polygenic traits relies on ever larger sample sizes and harmonized phenotype definitions and analysis techniques (50). As previously shown for depression (45), our study suggests that heterogeneity of phenotype definitions decreases the ability to identify genetic risk profiles that contribute to clinically relevant BD phenotypes. In addition to phenotype definitions, other factors that may limit cross-cohort comparability of the assessed phenotypes include demographic differences, cultural differences in diagnostic practice, and the uncertainty and measurement errors associated with recall bias, limited interrater reliability, and differences across health care systems (43,51–53). In our study, we performed both SNV-level and risk score associations using a structured meta-analysis, which mitigates some of the noise introduced by phenotypic heterogeneity. However, we are unable to account for differences in underlying genetic etiology of the phenotypes across cohorts. This heterogeneity is an important limitation of our study and should be considered in future phenotype analyses.

Our phenotype analyses confirmed the clinical relevance of disease onset phenotypes in BD. Age at BD onset was associated with important illness severity indicators, such as suicidality, positive symptoms, and achieving a lower level of education, thereby replicating findings of previous studies (25,28,54). Furthermore, patients with a depressive BD onset had an increased probability of lifetime suicidality, while those with a (hypo)manic onset were more likely to experience delusions and manic episodes (55–57). Contrary to previous evidence in a US (but not a French) sample, we observed that an earlier onset was associated with fewer episodes per illness year (56). Of note, when not normalizing for the illness duration, the age at onset was positively correlated with the number of episodes.

### 4.1. Conclusion

Phenotypes of BD onset are clinically important trait measures that represent the well-known clinical and biological heterogeneity of this severe psychiatric disorder. Genetic analysis of age and polarity at onset may lead to a better understanding of the biological risk factors underlying mental illness and support clinical assessment and prediction. Our study provides significant evidence of a genetic contribution to age and polarity at BD onset but also demonstrates the need for systematic harmonization of clinical data on BD onset in future studies. The coordinated efforts of international consortia and our growing ability to harness the rich phenotypic information stored in electronic health records will provide an unprecedented pool of phenotypic information on an increasingly diverse patient population (58–60). We expect that future genetic studies of age and polarity at onset will make major contributions toward our understanding of the etiology of BD and other psychiatric disorders.

## Supporting information

Ethics Overview

## Data Availability

Summary statistics for our meta-analysis of the GWAS cohort samples will be available through the PGC (https://www.med.unc.edu/pgc/download-results/).

## 5. Author contributions

Concept and design: Kalman, Olde Loohuis, Vreeker, Andlauer, Schulze, and Ophoff. Analysis and interpretation of data: Kalman, Olde Loohuis, Vreeker, Andlauer

Drafting of the manuscript: Kalman, Olde Loohuis, Vreeker, Andlauer

Supervision, and critical revision of the manuscript: Schulze, Ophoff, McMahon, Smoller, Alda.

All other authors provided data, contributed ideas and suggestions for analyses, interpreted results and revised the final manuscript.

## 6. Acknowledgments

The authors thank Jacquie Klesing, Board-certified Editor in the Life Sciences (ELS), for editing assistance with the manuscript.

*BOMA-Australia* sample: We thank Gin Mahli, Colleen Loo, and Micheal Breaskpear for their contribution to clinical assessments of a subset of patients and also Andrew Frankland for his work in collating clinical record data.

WTCCC sample: This study makes use of data generated by the Wellcome Trust Case-Control Consortium. A full list of the investigators who contributed to the generation of the data is available from www.wtccc.org.uk.

French sample: We thank the psychiatrists and psychologists who participated in the clinical assessment of patients in France (C. Henry, S. Gard, JP Kahn, L Zanouy, RF Cohen and O. Wajsbrot-Elgrabli) and thank the patients for their participation.

## 7. Funding

Loes M. Olde Loohuis: NIH K99/R00 MH116115.

Eli Stahl: NIH U01MH109536; E.S. is now employed by the Regeneron Genetics Center.

Andrew McQuillin: Medical Research Council, Grant/Award Numbers: G0500791, G0701007, G0801038, G1000708, G9623693N; Stanley Center for Psychiatric Research at the Broad Institute

Douglas Ruderfer: R01MH116269

Maria Grigoroiu-Serbanescu: UEFISCDI, Romania, several grants

Tim B Bigdeli: NIH MH085548, MH085542, MH104564

Fabian Streit: BMBF grant 01EW1810 ERA-Net Neuron “Synschiz”, BMBF grant

01ZX1614G e:Med Integrament

Mark J Adams: Wellcome Trust 104036/Z/14/Z, MRC MC_PC_17209

Rolf Adolfsson: Swedish Research Council (2009-33891-68296-196) and the Swedish Federal Government under the LUA/ALF agreement (ALF; RV-161691)

Ole A. Andreassen: Norwegian Research Council, KG Jebsen Stiftelsen, South-Eastern Norway Health Authority

Ceylan Balaban: BMBF “BipoLife” Subproject TPP1

Frank Bellivier: INSERM (Institut National de la Sante et de la Recherche Medicale - C0829), AP-HP (Assistance Publique des Hopitaux de Paris - RBM0436), Fondation FondaMental (RTRS Sante Mentale), Labex Bio-PSY (Investissements dAvenir program managed by the ANR under reference ANR-11-IDEX-0004-02).

Antonio Benabarre: Thanks the support of the Spanish Ministry of Science and Innovation (PI17/01122)

Wade Berrettini: R01 MH078156

Evelyn J Bromet: NIH MH085548, MH085542, MH104564

Sven Cichon: European Union Horizon 2020 Research and Innovation Programme (grant 785907 (HBP SGA2)), BMBF grant 01ZX1314Ae:Med Integrament, Swiss National Science Foundation (SNSF) grant 156791

William Coryell: R01 MH078154

Nick Craddock: Wellcome Trust (grant #078901)

David Craig: R01 MH078159

Paul E. Croarkin: National Institue of Mental Health (NIMH) R01 MH113700

Udo Dannlowski: This work was funded by the German Research Foundation (DFG, grant FOR2107 DA1151/5-1 and DA1151/5-2 to UD; SFB-TRR58, Projects C09 and Z02 to UD) and the Interdisciplinary Center for Clinical Research (IZKF) of the medical faculty of Munster (grant Dan3/012/17 to UD).

Franziska Degenhardt: German Federal Ministry of Education and Research (BMBF) within the e:Med programme (grant COMMITMENT) and the EU COST (European Cooperation in Science and Technology) programme (COST Action EnGagE CA17130).

Bruno Etain: INSERM (Institut National de la Sante et de la Recherche Medicale - C0829), AP-HP (Assistance Publique des Hopitaux de Paris - RBM0436), Fondation FondaMental (RTRS Sante Mentale), Labex Bio-PSY (Investissements dAvenir program managed by the ANR under reference ANR-11-IDEX-0004-02).

Ayman H Fanous: NIH MH085548, MH085542, MH104564

Janice M. Fullerton: National Health and Medical Research Council (Australia) grants 1037196,1063960, 1066177; and The Janette Mary ONeil Research Fellowship

Julie Garnham: Canadian Institutes of Health Research (grant #166098); Dalhousie Medical Research Foundation, Genome Atlantic, Lindsay family fund

Elliot Gershon: R01 MH078153

Tim Hahn: TH was supported by the German Research Foundation (DFG grants HA7070/2-2, HA7070/3, HA7070/4) and the Interdisciplinary Center for Clinical Research (IZKF) of the medical faculty of Munster (MzH3/020/20).

Stephane Jamain: INSERM (Institut National de la Sante et de la Recherche Medicale - C0829), AP-HP (Assistance Publique des Hopitaux de Paris - RBM0436), Fondation FondaMental (RTRS Sante Mentale), Labex Bio-PSY (Investissements dAvenir program managed by the ANR under reference ANR-11-IDEX-0004-02).

Esther Jimenez: EJ thanks the support of the Spanish Ministry of Science and Innovation (PI15/00283, PI18/00805) integrated into the Plan Nacional de I+D+I and co-financed by the ISCIII-Subdireccin General de Evaluacin and the Fondo Europeo de Desarrollo Regional (FEDER); the Instituto de Salud Carlos III; the CIBER of Mental Health (CIBERSAM); the Secretaria dUniversitats i Recerca del Departament dEconomia i Coneixement (2017 SGR 1365), the CERCA Programme, and the Departament de Salut de la Generalitat de Catalunya for the PERIS grant SLT006/17/00357. Ian Jones: Wellcome Trust (grant #078901) Lisa Jones: Wellcome Trust (grant #078901) John R. Kelsoe: R01 MH078151

Tilo Kircher: This work was funded by the German Research Foundation (DFG, grant FOR2107 KI588/14-1 and FOR2107 KI588/14-2 to TK)

George Kirov: Recriutment of the Bulgarian Trios was funded by the Janssen Research Foundation

James A Knowles: NIH MH085548, MH085542, MH104564

Thorsten Kranz: BMBF “BipoLife” Subproject TPP1

Trine Vik Lagerberg: Norwegian Research Council (grant #288542)

Mikael Landen: The Stanley Center for Psychiatric Research, Broad Institute from a grant from Stanley Medical Research Institute, the Swedish Research Council (2018-02653), the Swedish foundation for Strategic Research (KF10-0039), the Swedish Brain foundation (FO2020-0261), and the Swedish Federal Government under the LUA/ALF agreement (ALF 20170019, ALFGBG-716801).

William Lawson: R01 MH078161

Marion Leboyer: INSERM (Institut National de la Sante et de la Recherche Medicale - C0829), AP-HP (Assistance Publique des Hopitaux de Paris - RBM0436), Fondation FondaMental (RTRS Sante Mentale), Labex Bio-PSY (Investissements dAvenir program managed by the ANR under reference ANR-11-IDEX-0004-02).

Dolores MAlaspina: NIH MH085548, MH085542, MH104564

Melvin McInnis: R01 MH078162

Andrew M McIntosh: Wellcome Trust 104036/Z/14/Z, MRC MC_PC_17209

Helena Medeiros: NIH MH085548, MH085542, MH104564

Ingrid Melle: “Regional Health Authority South-Eastern Norway (grants # 2015088,2018093)”

Vihra Milanova: Recriutment of the Bulgarian Trios was funded by the Janssen Research Foundation

Philip B. Mitchell: National Health and Medical Research Council (Australia) grants 1037196, 1177991

John Nurnberger: R01 MH078152

Carlos Pato: NIH MH085548, MH085542, MH104564

Michele T Pato: NIH MH085548, MH085542, MH104564

James B. Potash: R01 MH078157

Mark H Rapaport: NIH MH085548, MH085542, MH104564

Andreas Reif: BMBF “BipoLife” Subproject TPP1

Marcella Rietschel: BMBF grant 01EW1810 ERA-Net Neuron “Synschiz”, BMBF grant 01EW1904 ERA-Net Neuron “Embed”, BMBF grant 01ZX01909A e:Med SysmedSUD, BMBF grant 01ZX1614G e:Med Integrament

Gloria Roberts: National Health and Medical Research Council (Australia) grants 1037196

Guy Rouleau: Canadian Institutes of Health Research (grant #166098)

William A. Scheftner: R01 MH078155

Peter R. Schofield: National Health and Medical Research Council (Australia) grants 1037196,1063960, 1176716

Janet L Sobell: NIH MH085548, MH085542, MH104564

John Strauss: Canadian Institutes of Health Research, MOP-172013 Gustavo Turecki: Canadian Institutes of Health Research (grant #166098)

Eduard Vieta: EV thanks the support of the Spanish Ministry of Science and Innovation (PI15/00283, PI18/00805) integrated into the Plan Nacional de I+D+I and co-financed by the ISCIII-Subdireccin General de Evaluacin and the Fondo Europeo de Desarrollo Regional (FEDER); the Instituto de Salud Carlos III; the CIBER of Mental Health (CIBERSAM); the Secretaria dUniversitats i Recerca del Departament dEconomia i Coneixement (2017 SGR 1365), the CERCA Programme, and the Departament de Salut de la Generalitat de Catalunya for the PERIS grant SLT006/17/00357.

John B. Vincent: Canadian Institutes of Health Research, MOP-172013 Jordan W Smoller: R01MH063445

Francis J. McMahon: Funded in part by the Intramural Research Program of the NIMH (ZIA MH002843)

Martin Alda: Canadian Institutes of Health Research (grant #166098); Dalhousie Medical Research Foundation, Genome Atlantic, Lindsay family fund

Till. F. M. Andlauer: German Federal Ministry of Education and Research (BMBF) through the DIFUTURE consortium of the Medical Informatics Initiative Germany (grant 01ZZ1804A) and the Integrated Network IntegraMent, under the auspices of the e:Med Programme (grant 01ZX1614J), as well as the European Unions Horizon 2020 Research and Innovation Programme (grant MultipleMS, EU RIA 733161)

The Collection of the Dutch cohort was funded through NIMH R01MH090553 awarded to Dr. Ophoff.

The collection of the Colombian samples was funded through NIMH R01 MH113078 awarded to Drs. Lopez-Jaramillo, Bearden and Freimer.

The collection of the *BOMA-Australia* sample (*bip_bmau_eur*) was supported by the Australian National Medical and Health Research Council (NHMRC) Program Grant 1037196 and Project Grants 1063960 and 1066177. DNA was extracted by Genetic Repositories Australia, an Enabling Facility that was supported by NHMRC Enabling Grant 401184. We gratefully acknowledge the Janette Mary ONeil Research Fellowship (to JMF). We acknowledge support from NHMRC Investigator Grants (Leadership 3) to PBM (1177991) and PRS (1176716).

Funding for the project was provided by the Wellcome Trust under award 076113, 085475 and 090355.

The work by the French group was supported by INSERM (Institut National de la Sante et de la Recherche Medicale - C0829), AP-HP (Assistance Publique des Hopitaux de Paris - RBM0436), the Fondation FondaMental (RTRS Sante Mentale), and the labex Bio-PSY (Investissements dAvenir program managed by the ANR under reference ANR-11-IDEX-0004-02).

## 8. Declaration of Interest

Amare T. Azmeraw: Dr Amare has received 2020-2022 NARSAD Young Investigator Grant from the Brain & Behaviour Research Foundation.

Ole A. Andreassen: Speaker’s honorarium Sunovion, Lundbeck. Consultant HealthLytix

Bernhard Baune: Honoraria: Lundbeck, Janssen, LivaNova, Servier

Carrie Bearden: Novartis Scientific Advisory Board

Clark Scott: Honoraria and Investigator Initiated project funding from Jannsen-Cilag Australia, Lundbeck-Otsuka Australia

J. Raymond DePaulo: owns stock in CVS Health; JRD was unpaid consultant for Myriad Neuroscience 2017 & 2019

Michael C. O’
sDonovan: Research unrelated to this manuscript supported by a collaborative research grant from Takeda Pharmaceuticals

Bruno Etain: Honoraria for Sanofi

Mark A. Frye: Grant Support Assurex Health, Mayo Foundation, Medibio Consultant (Mayo) Actify Neurotherapies, Allergan, Intra-Cellular Therapies, Inc., Janssen, Myriad, Neuralstem Inc., Sanofi, Takeda, Teva Pharmaceuticals

Per Hoffmann: Employee of Life&Brain GmbH, Member of the Scientific Advisory Board of HMG Systems Engeneering GmbH

Mikael Landen: Speaker’s honoraria Lundbeck pharmaceuticals

Andrew M McIntosh: Research funding from The Sackler Trust, speaker fees from Illumina and Janssen

Philip B. Mitchell: Remuneration for lectures in China on bipolar disorder research by Sanofi (Hangzhou)

John Nurnberger: Investigator for Janssen

Benjamin M. Neale: Is a member of the scientific advisory board at Deep Genomics and RBNC Therapeutics. A consultant for Camp4 Therapeutics, Takeda Pharmaceutical and Biogen.

Andreas Reif: Speaker’s honoraria / Advisory boards: Janssen, Shire/Takeda, Medice, SAGE and Servier

Eli Stahl: now employed by the Regeneron Genetics Center.

Kato Tadafumi: Honoraria: Kyowa Hakko Kirin Co., Ltd., Eli Lilly Japan K.K., Otsuka Pharmaceutical Co., Ltd., GlaxoSmithKline K.K., Taisho Pharma Co., Ltd., Taisho Pharmaceutical Co., Ltd., Taisho Toyama Pharmaceutical Co., Ltd., Dainippon Sumitomo Pharma Co., Ltd., Meiji Seika Pharma Co., Ltd., Pfizer Japan Inc., Mochida Pharmaceutical Co., Ltd., Shionogi & Co., Ltd., Janssen Pharmaceutical K.K., Janssen Asia Pacific, Yoshitomiyakuhin, Astellas Pharma Inc., Nippon Boehringer Ingelheim Co. Ltd., MSD K.K., Kyowa Pharmaceutical Industry Co., Ltd., Takeda Pharmaceutical Co., Ltd., Mitsubishi Tanabe Pharma Corporation, Eisai Co., Ltd. Grants: Takeda Pharmaceutical Co., Ltd., Dainippon Sumitomo Pharma Co., Ltd., Otsuka Pharmaceutical Co., Ltd., Shionogi & Co., Ltd., Eisai Co., Ltd., Mitsubishi Tanabe Pharma Corporation.

Eduard Vieta: Has received grants and served as consultant, advisor or CME speaker for the following entities: AB-Biotics, Abbott, Allergan, Angelini, Dainippon Sumitomo Pharma, Ferrer, Gedeon Richter, Janssen, Lundbeck, Otsuka, Sage, Sanofi-Aventis, Sunovion, and Takeda.

<u>None of the other Authors reported any biomedical financial interests or potential conflicts of interest.</u>

## 9. Availability of results

Summary statistics for our meta-analysis of the GWAS cohort samples are available through the PGC (https://www.med.unc.edu/pgc/download-results/).

