## Supplementary Material for "Characterization of Age and Polarity at Onset in Bipolar Disorder"

### Description of the individual study samples

The list below provides a detailed description (based on the original publications (1,2)) of all cohorts that were part of the present study.

#### 1. Discovery samples

***ume4 | Sweden***

Clinical characterization of the patients included the Mini-International Neuropsychiatric Interview (MINI), Diagnostic Interview for Genetic Studies (DIGS), Family Interview for Genetic Studies (FIGS), and Schedules for Clinical Assessment in Neuropsychiatry (SCAN). The final diagnoses were made according to the Diagnostic and Statistical Manual of Mental Disorders, fourth edition, text revision (DSM-IV-TR) and determined by consensus of 2 research psychiatrists.

***hal2 | Canada***

The case samples were recruited from patients longitudinally followed at specialty mood disorders clinics in Halifax and Ottawa (Canada). Cases were interviewed in a blind fashion with the Schedule of Affective Disorders and Schizophrenia-Lifetime version (SADS-L), and consensus diagnoses were made according to the Diagnostic and Statistical Manual of Mental Disorders, fourth edition (DSM-IV) and Research Diagnostic Criteria (RDC). Protocols and procedures were approved by the local ethics committees, and written informed consent was obtained from all patients before participation in the study.

***top7 | Norway***

In the TOP study (Thematically Organized Psychosis research), patients of European ancestry who were born in Norway were recruited from psychiatric hospitals in the Oslo region. Patients were diagnosed according to the Structured Clinical Interview (SCID) for DSM-IV. All participants provided written informed consent, and the Norwegian Scientific-Ethical Committee and the Norwegian Data Protection Agency approved the protocol.

***top8 | Norway***

The TOP8 bipolar disorder (BD) patients were recruited in the same way as the top7 cohort described above and recruited from hospitals across Norway.

***may1 | USA***

Cases of BD were drawn from the Mayo Clinic Bipolar Biobank. Enrolment sites included Mayo Clinic, Rochester, Minnesota; Lindner Center of HOPE/University of Cincinnati College of Medicine, Cincinnati, Ohio; and the University of Minnesota, Minneapolis, Minnesota. Enrolment at each site was approved by the local institutional review board (IRB), and all participants had consented to use of their data for future genetic studies. Participants were identified through routine clinical appointments, among inpatients in mood disorder units, and by recruitment advertising. They were required to be between 18 and 80 years old, be able to speak English and provide informed consent and have DSM-IV-TR diagnostic confirmation of BD-I or BD-II or schizoaffective bipolar disorder, as determined with the SCID.

***edi1 | UK***

This sample comprised Caucasian individuals contacted through the inpatient and outpatient services of hospitals in southeast Scotland. A BD-I diagnosis was based on an interview with the patient with the SADS-L, supplemented by case note review and frequently by information from medical staff, relatives, and caregivers. Final diagnoses, which were based on DSM-IV criteria, were reached by consensus between 2 trained psychiatrists. The study was approved by the Multi-Centre Research Ethics Committee for Scotland, and patients gave written informed consent for the collection of DNA samples for use in genetic studies.

***ageu |Sweden***

The patients with BD were identified in the Swedish National Quality Register for Bipolar Disorders (BipoläR) and the Swedish National Patient Register (with a validated algorithm that required at least 2 hospitalizations with a BD diagnosis). A confirmatory telephone interview with a diagnostic review was conducted. Additional patients were recruited from the St. Göran Bipolar Project (Affective Center at Northern Stockholm Psychiatry Clinic, Sweden), which enrolled new and ongoing patients diagnosed with BD by using structured clinical interviews. Diagnoses were made according to the DSM-IV (BipoläR and St. Göran Bipolar Project) and ICD-10 criteria (National Patient Register). All recruitment procedures were approved by the Regional EthicaBl Committees in Sweden.

***auom | Australia***

For the current analysis, datasets from the following study sites were combined and analyzed together:

*Adelaide subsample (n = 58):* Patients were collected as part of "The Cognitive Function and Mood Study (CoFaM-Study),” which was conducted at the Discipline of Psychiatry, University of Adelaide, Australia. Patients were diagnosed with the MINI diagnostic interview version 6.0. These diagnoses were compared with medical records and, for consistency reasons, the final clinical diagnosis was made following to DSM-IV criteria. The study was approved by ethics committees of the University of Adelaide and the Royal Adelaide Hospital. All participants provided written informed consent before study procedures were performed.

*Sydney subsample (n = 27):* Patients were recruited at the Mood Disorder Unit, Prince of Wales Hospital, in Sydney. All patients received a lifetime diagnosis of BD according to the DSM-IV criteria on the basis of a consensus best-estimate procedure and structured diagnostic interviews with the DIGS, FIGS, and SCID. Study protocols were reviewed and approved in advance by the IRBs of the participating institutions. All patients provided written informed consent.

***euo2 | Italy***

Patients were recruited among outpatients attending the Department of Psychiatry at the University of Campania “Luigi Vanvitelli” in Naples. All patients received a lifetime diagnosis of BD according to the DSM-IV criteria on the basis of a consensus best-estimate procedure that considered all available information, including the Structured Clinical Interview for DSM-IV Disorders (SCID-I)–Patient Edition (SCID-IP), medical records, and a routine clinical interview. In addition, psychopathological rating scales and the retrospective chart of the National Institute of Mental Health Life Chart Method were used for detailed and longitudinal assessment of clinical aspects of BD. The study was approved by the local IRB. All participants provided written informed consent.

***euoI | Italy***

The sample comprised 181 unrelated patients with BD I. All patients were of Sardinian ancestry for at least 3 generations. They were recruited at the outpatient unit (Lithium Clinic) of the Clinical Psychopharmacology Center at the Department of Biomedical Science, Section of Neuroscience & Clinical Pharmacology, University of Cagliari, Cagliari, Italy, and Unit of Clinical Pharmacology, University Hospital Agency of Cagliari, Cagliari, Italy. Lifetime consensus diagnoses according to RDC were made by trained clinical psychopharmacologists on the basis of data from a personal semi-structured interview and a systematic review of patients’ medical records. Informed written consent to participate in the study was obtained from all patients. The study was approved by the local ethics committee.

***euoR | Austria, Czech Republic, France, Germany, Romania, Spain, Switzerland, Sweden***

For the current analysis, datasets from the following study sites were combined and analyzed together:

*Austrian subsample (n = 35):* Patients were recruited at the Medical University of Graz, Department of Psychiatry and Psychotherapeutic Medicine. All patients received a lifetime diagnosis of BD according to the DSM-IV criteria on the basis of a consensus best-estimate procedure that considered all available information, including structured diagnostic interviews with the SCID, medical records, and personal medical history. Study protocols were reviewed and approved in advance by the IRBs of the participating institutions. All participants provided written informed consent.

*Czech subsample (n = 45):* Unrelated patients were recruited in in- and outpatient units at the Prague Psychiatric Center, Psychiatric Hospital Bohnice, Psychiatric Clinic, Czech Republic. Diagnoses were made on the basis of either a SADS-L interview or on an unstructured clinical interview modified from SADS-L by using RDC criteria. All patients signed an informed consent form approved by the IRBs of the Prague Psychiatric Center.

*French subsample (n = 46):* The research sample comprised participants recruited between 1995 and 2008 from 3 university-affiliated departments of psychiatry (Paris, Bordeaux, and Nancy) in France. The inclusion criteria were that the individual (a) was aged 18 years or older; (b) had a mood disorder that met DSM-IV criteria for BD-I or BD-II or BD not otherwise specified (BDNOS); (c) currently met criteria for euthymia, which was operationalized as scores below 5 on both the Montgomery-Åsberg Depression Rating Scale and the Bech Mania Rating Scale; and (d) was willing and able to give written informed consent. Study protocols were reviewed and approved by the IRBs of the participating institutions. Patients meeting the above inclusion criteria were assessed by psychiatrists trained in the use of the French version of the DIGS.

*German subsample (n = 71):* Patients were recruited from consecutive admissions to psychiatric inpatient units at the University Hospital Würzburg. All patients received a lifetime diagnosis of BD according to the DSM-IV criteria on the basis of a consensus best-estimate procedure that considered all available information, including semi-structured diagnostic interviews using the Association for Methodology and Documentation in Psychiatry (AMDP), medical records, and the family history method. In addition, the Operational Criteria Checklist for Psychotic Illness (OPCRIT) was used for detailed polydiagnostic documentation of symptoms. Study protocols were reviewed and approved in advance by the IRBs of the participating institutions. All patients provided written informed consent.

*Romanian subsample (n = 8):* Patients who had taken lithium for at least 2 years (lithium treatment response evaluated with the Retrospective Criteria of Long-Term Treatment Response in Research Subjects with Bipolar Disorder [Alda scale]) were recruited from consecutive admissions to the Obregia Clinical Psychiatric Hospital, Bucharest, Romania. Patients were interviewed with the DIGS and FIGS. Information was also obtained from medical records and close relatives. The diagnosis of BD-I was assigned according to DSM-IV criteria by using the best estimate procedure. Patients were included in the sample if they had at least 2 documented hospitalizations for illness episodes (1 manic/mixed and 1 depressive or 2 manic episodes). All participants provided written informed consent. The study was performed in accordance with the ethical principles of the Declaration of Helsinki.

*Spanish subsample (n = 73):* Cases were recruited from the [Bipolar Disorder](https://www-sciencedirect-com.emedien.ub.uni-muenchen.de/topics/medicine-and-dentistry/bipolar-disorder) Program of the Hospital Clinic of Barcelona and [Mental Health Services](https://www-sciencedirect-com.emedien.ub.uni-muenchen.de/topics/medicine-and-dentistry/mental-health-service) from Oviedo, under the umbrella of the Spanish Research Network on Mental Health (CIBERSAM). Participants were selected only if they fulfilled the following inclusion criteria: (i) met DSM-IV-TR criteria for BD-I or -II, (ii) age over 18 years, (iii) met criteria for euthymia for at least 3 months before inclusion, assessed by the Hamilton Depression Rating Scale (HDRS) and the Young Mania Rating Scale (YMRS), and (iv) provided both written and verbal informed consent. Exclusion criteria were as follows: (i) intelligence quotient (IQ) lower than 70, (ii) the presence of any medical condition affecting neuropsychological performance, and (iii) [electroconvulsive therapy](https://www-sciencedirect-com.emedien.ub.uni-muenchen.de/topics/medicine-and-dentistry/electroconvulsive-therapy) within the past year. The study was approved by each institution’s ethics committees and was performed in accordance with the ethical principles of the Declaration of Helsinki.

*Swedish subsample (n = 80):* The patients with BD were identified in the Swedish National Quality Register for Bipolar Disorders (BipoläR) and the Swedish National Patient Register (with a validated algorithm that required at least 2 hospitalizations with a BD diagnosis). A confirmatory telephone interview with a diagnostic review was conducted. Additional patients were recruited from the St. Göran Bipolar Project (Affective Center at Northern Stockholm Psychiatry Clinic, Sweden), which enrolled new and ongoing patients diagnosed with BD by using structured clinical interviews. Diagnoses were made according to the DSM-IV (BipoläR and St. Göran Bipolar Project) and ICD-10 criteria (National Patient Register). All recruitment procedures were approved by the Regional Ethical Committees in Sweden.

*Swiss subsample (n = 52):* Patients with BD were recruited in the specialized outpatient unit for mood disorders at the Division of Psychiatric Specialties of the Department of Psychiatry in Geneva. Patients are referred to this unit by psychiatrists or general practitioners for diagnostic assessment and care. Individuals diagnosed by a trained psychiatrist or clinical psychologist with a DSM-IV diagnosis of BD-I or -II were included in this study. Clinical and anamnestic data (medical histories, family history, onset of the disorder, and previous treatments) were collected during the interview. Patients with BD were evaluated with the French version of the DIGS or the French version of the SCID (SCID I, version 2.0). They were also evaluated for comorbid Axis I disorders with the DIGS. Treatment response to lithium was evaluated with the Alda scale. All patients completed self-report questionnaires, including the Barrat Impulsiveness Scale (BIS-10), State Anger Expression Inventory (STAXI), Beck Hopelessness Scale (BHS), Childhood Trauma Questionnaire (CTQ), Brown-Goodwin Aggression Scale (BGA) and the Geneva Suicide History Form. All patients provided written informed consent.

***h66x| Germany, Poland***

For the current analysis, datasets from the following study sites were combined and analyzed together:

*Polish subsample (n = 88):* Patients were recruited at the Department of Psychiatry, Poznan University of Medical Sciences, Poznan, Poland. All patients received a lifetime diagnosis of BD according to the DSM-IV criteria on the basis of a consensus best-estimate procedure and structured diagnostic interviews with the SCID. Study protocols were reviewed and approved in advance by the IRBs of the participating institutions. All subjects provided written informed consent.

*German subsample (n = 4):* Patients were recruited from consecutive admissions to the in- and outpatient units of the Department of Psychiatry and Psychotherapy at the Universities of Dresden and Berlin (Charité), Germany. DSM-IV lifetime diagnoses of BD-I were assigned on the basis of a consensus best-estimate procedure that considered all available information, including a structured interview with the SCID and medical records. Study protocols were reviewed and approved in advance by the IRBs of the participating institutions. All patients provided written informed consent.

***naom | USA***

For the current analysis, datasets from the following study sites were combined and analyzed together:

*Baltimore subsample (n = 11):* Patients were recruited at the Johns Hopkins Hospital in Baltimore, Maryland, USA. All patients received a lifetime diagnosis of BD-I according to the DSM-IV criteria on the basis of a consensus best-estimate procedure that considered all available information, including structured diagnostic interviews with the Diagnostic Interview for Genetic Studies, medical records, and the family history method. Study protocols were reviewed and approved in advance by the IRB of the Johns Hopkins Hospital. All patients provided written informed consent.

Iowa City subsample (n = 13): Patients were recruited at the University of Iowa Hospitals and Clinics in Iowa City, Iowa, USA. All patients received a lifetime diagnosis of BD-I according to the DSM-IV criteria on the basis of a consensus best-estimate procedure that considered all available information, including structured diagnostic interviews with the DIGS, medical records, and the family history method. Study protocols were reviewed and approved in advance by the IRB of the University of Iowa, Carver College of Medicine. All patients provided written informed consent.

NIMH subsample (n = 17): Patients were recruited in the same way as those in the gain cohort. Study protocols were reviewed and approved in advance by the IRBs of the participating institutions. All patients provided written informed consent.

Rochester subsample (n = 26): Cases of BD were drawn from the Mayo Clinic Bipolar Biobank. Enrolment sites included Mayo Clinic, Rochester, Minnesota; Lindner Center of HOPE/University of Cincinnati College of Medicine, Cincinnati, Ohio; and the University of Minnesota, Minneapolis, Minnesota. Enrolment at each site was approved by the local IRB, and all participants had consented to the use of their data in future genetic studies. Participants were identified through routine clinical appointments, among inpatients admitted in mood disorder units, and by recruitment advertising. Participants were required to be between 18 and 80 years old and be able to speak English and provide informed consent and have DSM-IV-TR diagnostic confirmation of BD-I or -II or schizoaffective bipolar disorder, as determined with the SCID.

San Diego subsample (n = 92): Patients were recruited from individuals at the University of California, San Diego, as described for the gain samples below.

***dub1 | Ireland***

Samples were collected as part of a larger study on the genetics of psychotic disorders in the Republic of Ireland, under protocols approved by the relevant IRBs and with written informed consent that permitted repository use. Patients were recruited from hospitals and community psychiatric facilities in Ireland by a psychiatrist or psychiatric nurse trained in using the SCID. Diagnosis was based on the structured interview and supplemented by case note review and collateral history, where available. All diagnoses were reviewed by an independent reviewer.

***wtccc | United Kingdom***

Patients were all over the age of 17 years, living in the UK, and of European descent. Recruitment was undertaken throughout the UK and included individuals who had been in contact with mental health services and had a lifetime history of high mood. After providing written informed consent, participants were interviewed by a trained psychologist or psychiatrist with a semi-structured lifetime diagnostic psychiatric interview (SCAN) and available psychiatric medical records were reviewed. On the basis of all available data, best-estimate life-time diagnoses were made according to the RDC. In the current study, we included cases with a lifetime diagnosis of RDC BD-I, BD-II, or schizoaffective disorder, bipolar type. All patients were recruited under protocols approved by the appropriate IRBs and gave written informed consent.

***bmau | Australia***

Patients were recruited at the Mood Disorder Unit, Prince of Wales Hospital in Sydney. All patients received a lifetime diagnosis of BD according to the DSM-IV criteria on the basis of a consensus best-estimate procedure and structured diagnostic interviews with the DIGS, FIGS, and SCID. Study protocols were reviewed and approved in advance by the IRBs of the participating institutions. All patients provided written informed consent.

***rom3 | Romania***

Patients with BD-I (n = 233) were recruited from consecutive admissions to the Obregia Clinical Psychiatric Hospital, Bucharest, Romania. Patients were interviewed with the DIGS and FIGS. Information was also obtained from medical records and close relatives. The diagnosis of BD-I was assigned according to DSM-IV criteria on the basis of the best estimate procedure. All patients had at least 2 hospitalizations for illness episodes. All participants provided written informed consent. The study was performed in accordance with the ethical principles of the Declaration of Helsinki.

***bmpo | Poland***

Patients were recruited at the Department of Psychiatry, Poznan University of Medical Sciences, Poznan, Poland. All patients received a lifetime diagnosis of BD according to the DSM-IV criteria on the basis of a consensus best-estimate procedure and structured diagnostic interviews with the SCID. Study protocols were reviewed and approved in advance by the IRBs of the participating institutions. All patients provided written informed consent.

***gain | USA***

*Genetic Association Information Network (GAIN)/ The Bipolar Genome Study (BiGS)* The BD sample was collected under the auspices of the NIMH Genetics Initiative for BD (<http://zork.wustl.edu/nimh/>), genotyped as part of GAIN and analyzed as part of a larger GWAS conducted by the BiGS consortium. Approximately half of the GAIN sample was collected as multiplex families or sib-pair families (waves 1-4), and the remainder was collected as individual cases (wave 5). Patients were recruited at 11 sites: Indiana University; John Hopkins University; the NIMH Intramural Research Program; Washington University at St. Louis; University of Pennsylvania; University of Chicago; Rush Medical School; University of Iowa; University of California, San Diego; University of California, San Francisco; and University of Michigan. All investigations were carried out after the review of protocols by the IRB at each participating institution. At all sites, potential patients were identified from screening admissions to local treatment facilities and through publicity programs or advocacy groups and evaluated with the DIGS, FIGS, and information from relatives and medical records. All information was reviewed through a best-estimate diagnostic procedure by 2 independent and non-interviewing clinicians and a best-estimate diagnosis was reached. In the event of a disagreement, a third review was performed to break the tie.

***tgco2 | USA***

Patients were recruited from individuals at the 11 US sites described for the GAIN sample as part of FAT2, FaST, BiGS, and TGEN cohorts. Eligible participants were aged 18 or older and met DSM-IV criteria for BD-I or BD-II by consensus diagnosis based on interviews with the Affective Disorders Evaluation (ADE) and MINI. All participants provided written informed consent, and the study protocol was approved by the IRB at each site. Collection of phenotypic data and DNA samples was supported by NIMH grants MH063445 (JW Smoller); MH067288 (PI: P Sklar), MH63420 (PI: V Nimgaonkar), and MH078151, MH92758 (PI: J. Kelsoe). The samples were independent of those included in the GAIN sample.

***butr | Bulgaria***

All patients were recruited in Bulgaria from psychiatric inpatient and outpatient services. Each patient had a history of hospitalization and was interviewed with an abbreviated version of the SCAN. Consensus best-estimate diagnoses were made by 2 researchers according to DSM-IV criteria. All participants gave written informed consent, and the study was approved by local ethics committees at the participating centers.

***swa2 | Sweden***

The patients with BD were identified in the Swedish National Quality Register for Bipolar Disorders (BipoläR) and the Swedish National Patient Register (with a validated algorithm that required at least two hospitalizations with a BD diagnosis). A confirmatory telephone interview with a diagnostic review was conducted. Additional patients were recruited from the St. Göran Bipolar Project (Affective Center at Northern Stockholm Psychiatry Clinic, Sweden), which enrolled new and ongoing patients diagnosed with BD with structured clinical interviews. Diagnoses were made according to the DSM-IV (BipoläR and St. Göran Bipolar Project) and ICD-10 criteria (National Patient Register). All recruitment procedures were approved by the Regional Ethical Committees in Sweden.

***fran | France***

Patients with BD-I or BD-II were recruited as part of a large study on the genetics of BD in France (Paris-Creteil, Bordeaux, Nancy) with a protocol approved by the relevant IRBs and with written informed consent. Patients were of French descent for more than 3 generations and were assessed by a trained psychiatrist or psychologist with structured interviews, supplemented by medical case notes, mood scales, and a self-rating questionnaire that assessed dimensions.

***uclo | United Kingdom***

The UCL sample comprised Caucasian individuals who were recruited by and received clinical diagnoses of BD-I from UK National Health Service (NHS) psychiatrists at interview on the basis of the criteria of the ICD-10. In addition, patients with BD were included only if both parents were of English, Irish, Welsh, or Scottish descent and if 3 out of 4 grandparents were of the same descent. All patients read an information sheet approved by the Metropolitan Medical Research Ethics Committee, which also approved the project for all NHS hospitals. Written informed consent was obtained from each patient.

***bonn | Germany***

Patients for the BOMA-Bipolar Study were recruited from consecutive admissions to the inpatient units of the Department of Psychiatry and Psychotherapy at the University of Bonn and the Central Institute for Mental Health in Mannheim, University of Heidelberg, Germany. DSM-IV lifetime diagnoses of BD-I were assigned on the basis of a consensus best-estimate procedure that considered all available information, including a structured interview with the SCID and SADS-L, medical records, and the family history method. In addition, the OPCRIT checklist was used for the detailed polydiagnostic documentation of symptoms. Study protocols were reviewed and approved in advance by the IRBs of the participating institutions. All patients provided written informed consent.

***bmg2 | Germany***

Patients were recruited from consecutive admissions to psychiatric inpatient units at the University Hospital Würzburg. All patients received a lifetime diagnosis of BD according to the DSM-IV criteria on the basis of a consensus best-estimate procedure that considered all available information, including semi-structured diagnostic interviews with the AMDP, medical records, and the family history method. In addition, the OPCRIT system was used for the detailed polydiagnostic documentation of symptoms. Study protocols were reviewed and approved in advance by the IRBs of the participating institutions. All patients provided written informed consent.

***bmg3 | Germany***

Patients were recruited at the Central Institute of Mental Health in Mannheim, University of Heidelberg, and other collaborating psychiatric hospitals in Germany. All patients received a lifetime diagnosis of BD according to the DSM-IV criteria on the basis of a consensus best-estimate procedure that considered all available information, including structured diagnostic interviews with the AMDP, Composite International Diagnostic Screener (CID-S), SADS-L, and/or SCID, medical records, and the family history method. In addition, the OPCRIT system was used for the detailed polydiagnostic documentation of symptoms. Study protocols were reviewed and approved in advance by the IRBs of the participating institutions. All patients provided written informed consent.

***bmsp | Spain***

Patients were recruited at the mental health departments of the following 5 centers in Andalusia, Spain: University Hospital Reina Sofia of Córdoba, Provincial Hospital of Jaen; Hospital of Jerez de la Frontera (Cádiz); Hospital of Puerto Real (Cádiz); Hospital Punta Europa of Algeciras (Cádiz); and Hospital Universitario San Cecilio (Granada). Diagnoses were made on the basis of the SADS-L, OPCRIT, a review of medical records, and interviews with first- and/or second-degree family members with the Family Informant Schedule and Criteria (FISC). Consensus best-estimate BD diagnoses were assigned by 2 or more independent senior psychiatrists and/or psychologists and according to the RDC and DSM-IV. Study protocols were reviewed and approved in advance by the IRBs of the participating institutions. All patients provided written informed consent.

***ucla | Netherlands***

The case sample consisted of in- and outpatients recruited through psychiatric hospitals and institutions throughout the Netherlands. Patients with DSM-IV BD, determined after interview with the SCID, were included in the analysis. Ethical approval was provided by UCLA and local ethics committees, and all participants gave written informed consent.

***usc2 | USA***

Genomic Psychiatry Consortium (GPC) patients were recruited via the University of Southern California healthcare system. Diagnoses were based on DSM-IV-TR criteria and were established with the OPCRIT on the basis of a combination of focused, direct interviews and data extracted from medical records.

***mich | USA***

The Pritzker Neuropsychiatric Disorders Research Consortium (NIMH/Pritzker) patients were from the NIMH Genetics Initiative Genetics Initiative Repository. Patients were diagnosed according to DSM-III or DSM-IV criteria by diagnostic interviews and/or medical record reviews. Cases with low confidence diagnoses were excluded. From each non-Ashkenazi European-origin family available from wave 1-5, 2 siblings with BD-I were included, when possible, and the patients was preferentially included, if possible (n = 946 individuals in 473 sibling pairs); otherwise, a single patient with BD-I was included (n = 184). The sibling pairs with BD were retained within the NIMH/Pritzker sample when individuals in more than 1 study were uniquely assigned to a study set.

***stp1 | USA***

The Systematic Treatment Enhancement Program for Bipolar Disorder (STEP-BD) was a 7-site, national US, longitudinal cohort study designed to examine the effectiveness of treatments and their impact on the course of BD. The study enrolled 4361 participants who met DSM-IV criteria for BD-I, BD-II, BDNOS, schizoaffective manic or bipolar type, or cyclothymic disorder on the basis of diagnostic interviews. From the parent study, 2089 individuals with BD-I or -II diagnoses who were over 18 years of age consented to the collection of blood samples for DNA. BD samples with a consensus diagnosis of BD-I were selected for inclusion in STEP1.

***m&m‘s | Germany, Austria***

PsyCourse subsample (n = 365): The samples form part of a multi-site German/Austrian longitudinal study ([www.psycourse.de](http://www.psycourse.de)).(3) Diagnoses were made according to DSM-IV. Study protocols were reviewed and approved in advance by the IRBs of the participating institutions. All patients provided written informed consent. The current analyses are based on the v3.1. version of the data set.

FOR2017 subsample (n = 88): In- and outpatients (aged 18-65) were recruited as part of an ongoing multi-center (Universities of Marburg and Münster, Germany) cohort study (DFG research group FOR2107, [www.for2107.de](http://www.for2107.de)). Trained psychologists conducted semi-structured interviews for DSM-IV axis I disorders (SCID-I). The study protocols were approved by the ethics committees of the Medical Schools of the Universities of Marburg and Münster, following the Declaration of Helsinki, and all participants provided written informed consent.

#### 2. Replication samples

***bdtrs | Germany***

The Bipolar Disorder Treatment Response Study (BP-TRS) comprises inpatients with BD and screened controls of Caucasian background. A trained psychologist or psychiatrist conducted a face-to-face interview with SCID or MINI 6.0 to ascertain the presence of BD according to DSM-IV criteria. Patients aged 18 years and older were included if a current or lifetime diagnosis of BD was determined in this structured diagnostic interview. Other assessments, including symptom ratings, psychiatric history, treatment history, and treatment response, were based on an interview by trained psychologists/psychiatrists. All patients provided written informed consent.

***ukwa1 | United Kingdom***

The UCL sample comprised Caucasian individuals who were recruited by and received clinical diagnoses of BD-I from UK National Health Service (NHS) psychiatrists at interview on the basis of the criteria of the ICD-10. In addition, patients with BD were included only if both parents were of English, Irish, Welsh, or Scottish descent and if 3 out of 4 grandparents were of the same descent. All volunteers read an information sheet approved by the Metropolitan Medical Research Ethics Committee, which also approved the project for all NHS hospitals. Written informed consent was obtained from each volunteer.

***dutch | Netherlands***

The case sample consisted of in- and outpatients recruited through psychiatric hospitals and institutions throughout the Netherlands. Patients with DSM-IV BD, determined after interview with the SCID, were included in the analysis. Ethical approval was provided by UCLA and local ethics committees, and all participants gave written informed consent.

***bmrom | Romania***

The sample *bmrom* (N = 225 BD-I cases) also included patients from the ConLiGen-Romania sample who did not overlap with the Romanian PGC2 sample bip_rom3_eur. Patients with BD-I were recruited from consecutive admissions in the Obregia Psychiatric Hospital of Bucharest, Romania. Patients were interviewed with the DIGS and FIGS. Information was also obtained from medical records and close relatives. The diagnosis of BD-I was assigned according to DSM-IV criteria by the best estimate procedure. Patients were included in the sample if they had at least 2 documented hospitalizations for illness episodes (1 manic/mixed and 1 depressive or 2 manic episodes). All participants provided written informed consent. The study was performed in accordance with the ethical principles of Declaration of Helsinki. Patients in the ConLiGen-Romania study were recruited in the same manner as the other patients in the ***bmrom*** sample and were required to have taken lithium for at least 2 years; lithium treatment response was evaluated with the Alda scale.

***jst5 | USA***

The study included unrelated patients with BD-I from 6 clinical trials (IDs: NCT00253162, NCT00257075, NCT00076115, NCT00299715, NCT00309699, and NCT00309686). Janssen Research & Development, LLC (formerly known as Johnson & Johnson Pharmaceutical Research & Development, LLC) recruited participants to assess the efficacy and safety of risperidone. Patients were diagnosed with BD according to DSM-IV-TR criteria. The diagnosis of BD was confirmed by the Schedule for Affective Disorders and Schizophrenia for School-Age Children-Present and Lifetime Version (K-SADS-PL) in NCT00076115, by the SCID in NCT00257075 and NCT00253162, and by the MINI in NCT00299715, NCT00309699, and NCT00309686. Additional detailed descriptions of these clinical trials can be found at ClinicalTrials.gov. Only patients of European ancestry were included in the current analysis.

***col1 | Colombia***

Patients with BD-I and -II were recruited as part of a larger cohort of patients with severe mental illness through psychiatric hospitals in the Paisa region of Colombia. Protocols and procedures were approved by the local and UCLA ethics committees, and written informed consent was obtained from all patients before participation in the study. Phenotyping included diagnostic interview (NetSCID-5, Spanish version), additional assessments of individual symptoms, and a neurocognitive battery.

### Supplementary Methods

#### Phenotype analyses: Statistical analysis

We analyzed the relationship of age at onset and polarity at onset with disease characteristics only in patients with BD-I. To optimize comparability between studies, we dichotomized the following variables: lifetime delusions, lifetime hallucinations, suicidal ideation, suicide attempt, educational attainment, current smoking, and living together with a partner (all variables were dichotomized as yes/no except for educational attainment, which was dichotomized as lower/higher; see Supplementary Table S9). We considered the number of manic and depressive episodes as continuous variables. However, to adjust for illness duration we calculated the frequency of episodes per year as: ((number of episodes) / (years of illness + 1)). Then, we rank-normalized these variables for analysis.

We analyzed the associations of age at onset and polarity at onset with dichotomous illness characteristics by logistic regression analysis. The various dichotomous illness characteristics were used as the outcome, and either age at onset or polarity at onset was used as the determinant. In addition, we analyzed the associations of age at onset and polarity at onset with the frequency of episodes per year by separate linear regression analyses. Sex was included as a covariate in all analyses. Regression analyses were performed in SPSS 25.0. Results from both datasets were then combined by a fixed-effects meta-analysis in R (package Metafor). We applied the Bonferroni-Holm method to correct for multiple testing.

#### Phenotype analyses: Secondary analysis

Since our finding that a later AAO was associated with a higher frequency of episodes per year of illness were contrary to previous findings in other studies (Etain *et al.*), we conducted secondary analyses in which we used the normalized age at onset and gender as predictors and the untransformed number of episodes (not controlled for years of illness) as outcome variables. The distribution of the residuals of these models did not follow a normal distribution. Therefore, these results should be interpreted with caution.

In the Dutch study, the association of the age at onset with less manic episodes was not significant (β=-0.41, SE=0.25, *p*=1.01×10^-1^). However, a later age at onset was significantly associated with less depressive episodes (β=-1.04, SE=0.41, *p*=1.03×10^-2^). In the German dataset, a later age at onset was significantly associated with less manic (β=-1.48, SE=0.46, *p*=1.35×10^-3^) and depressive episodes (β=-2.04, SE=0.56, *p*=3.72×10^-4^).

### Supplementary Figures

#### Supplementary Figure S1. Proportions of each category of polarity at onset by continent and definition of age at onset

Categories of polarity at onset (PAO): D, depression before mania/hypomania; M, mania/hypomania before depression; Mixed, mixed episodes or first manic and depressive episode in same year.

**A:** Proportions of PAO by dataset and continent

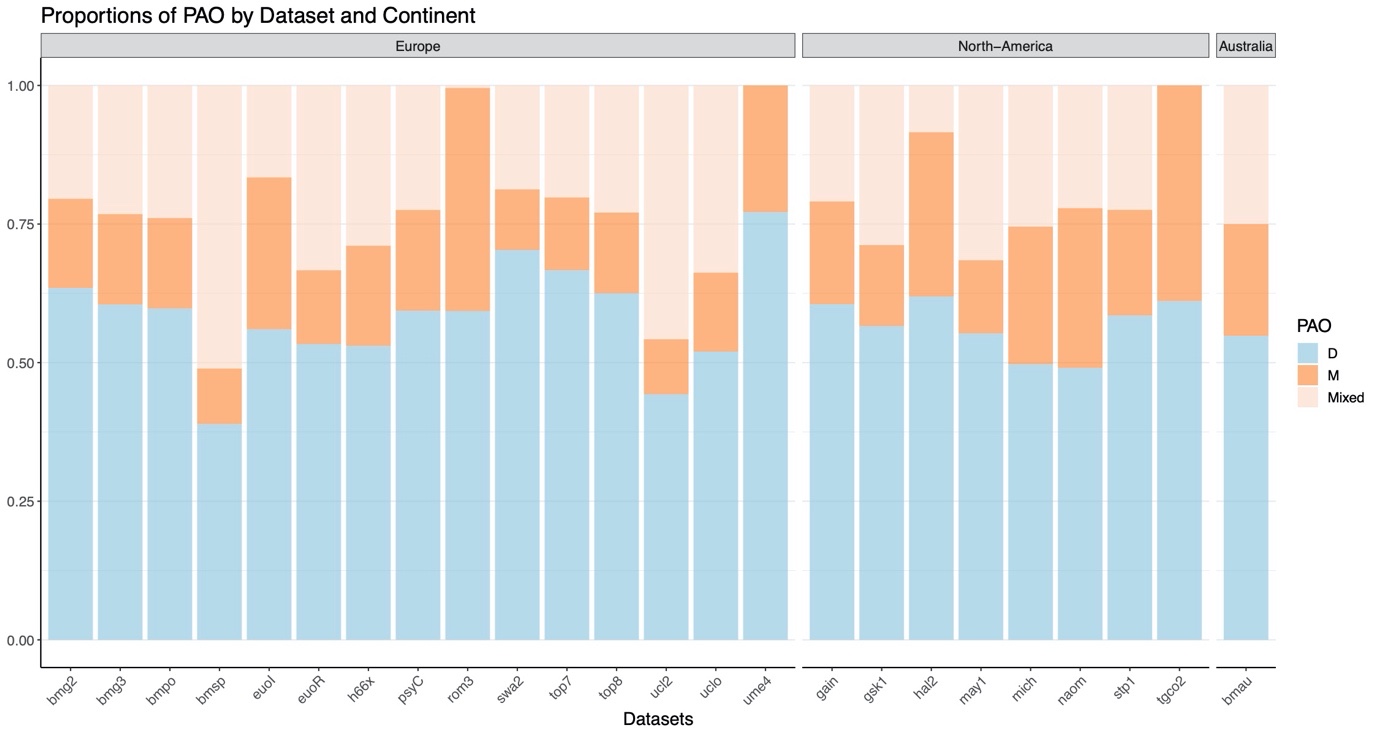

**B:** Proportions of PAO by dataset and definition of age at onset

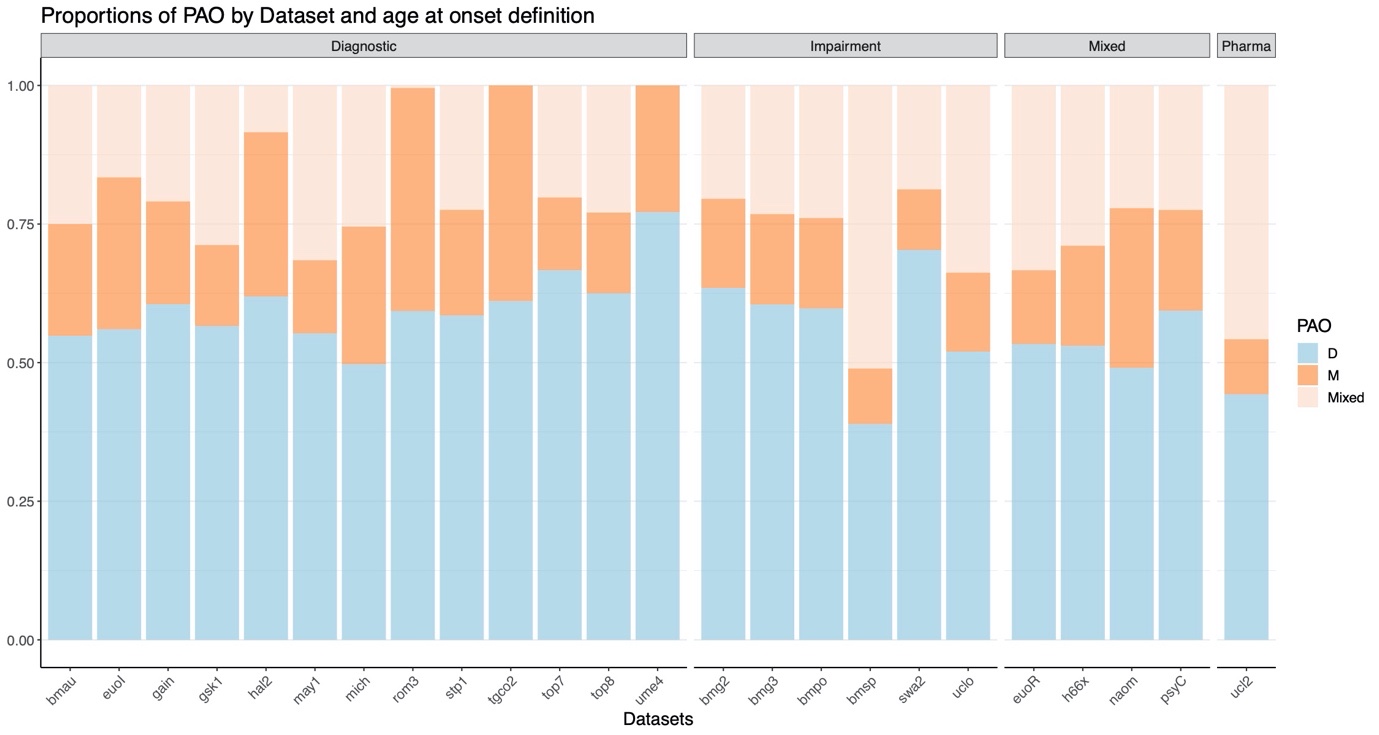

#### Supplementary Figure S2. Forest plots from the GWAS on age at onset

**A:** Forest plot of the results of the discovery-stage analysis of the top-associated variant rs1610275. The effect size beta is relative to the minor allele G. The color scheme corresponds to the colors used in Fig. 1

**B:** Forest plot of the results of the replication-stage analysis of variant rs1610275

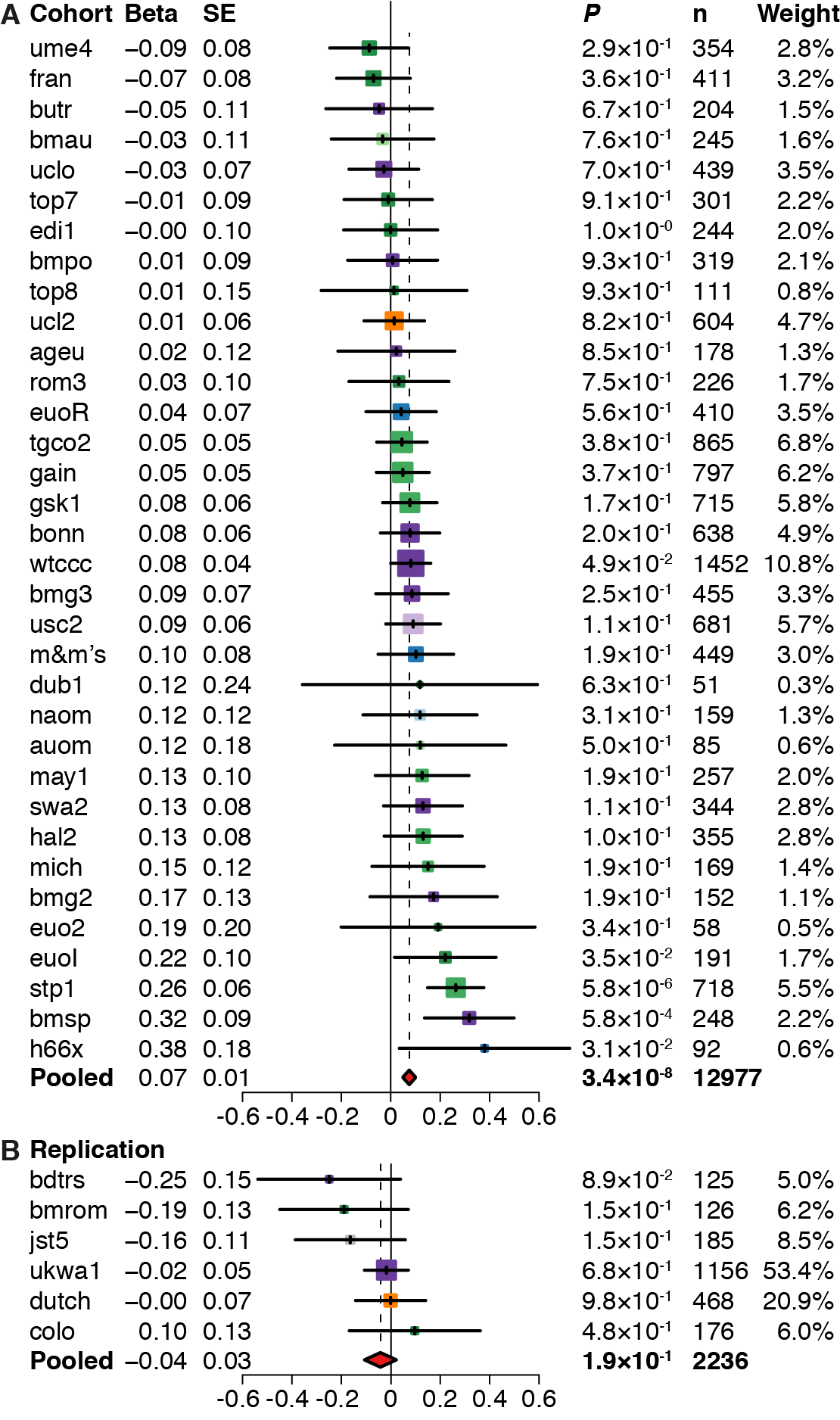

#### Supplementary Figure S3. Results of the GWAS on polarity at onset

**A:** Manhattan plot of the primary PAO (PAO-M/-X [n=2888] *vs.* PAO-D [n=3885]) GWAS

**B:** Manhattan plot of the secondary PAO (PAO-M [n=1350] *vs.* PAO-D [n=3599]) GWAS

Abbreviations: PAO-M, mania/hypomania before depression; PAO-X, mixed episodes or first manic and depressive episode in same year; PAO-D, depression before mania/hypomania.

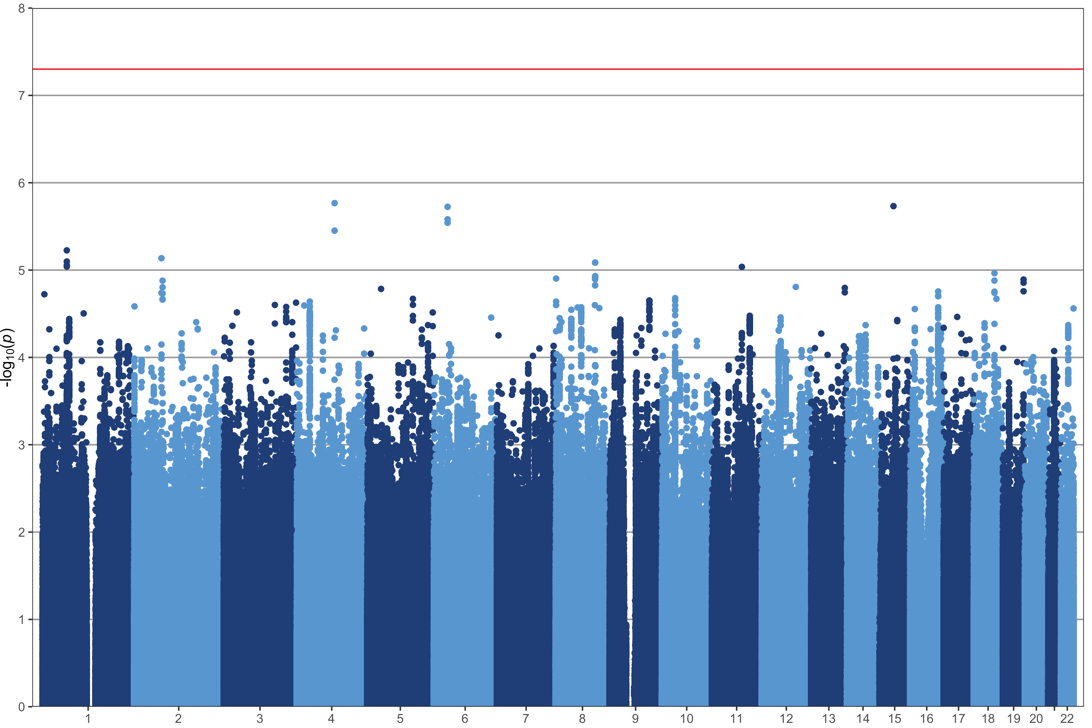

**A**

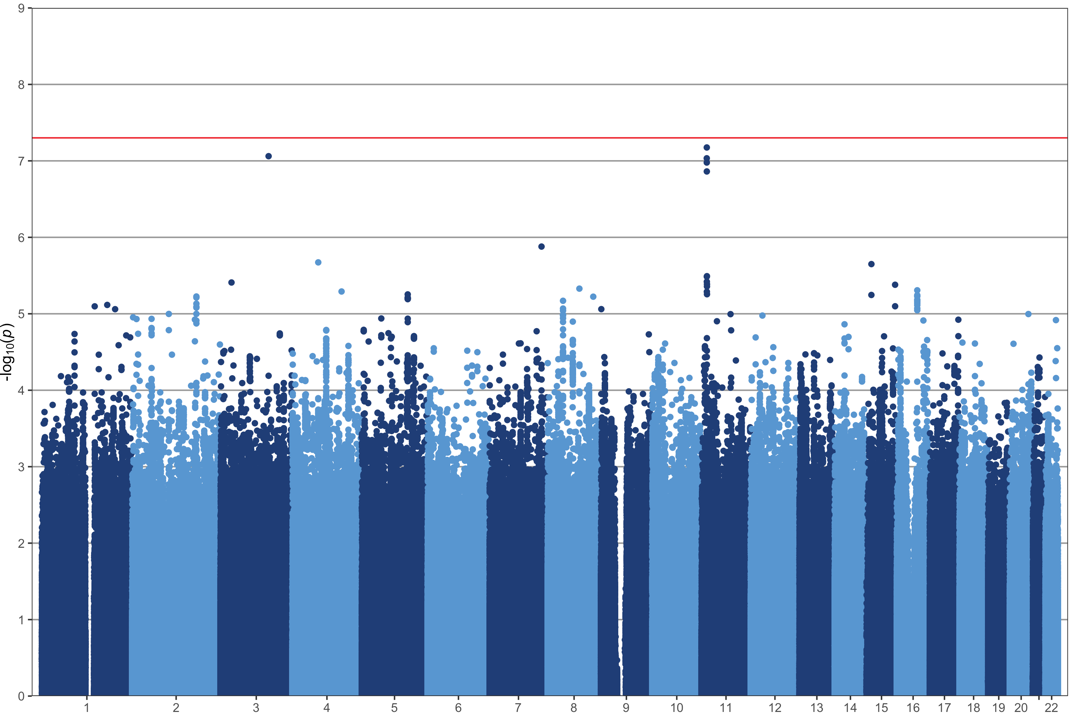

**B**

#### Supplementary Figure S4: Results from analyses of polygenic scores with the polarity of onset

**A:** Associations of polygenic scores (PGSs) with the polarity at onset (PAO-M and PAO-X *vs.* PAO-D). A higher odds ratio (OR) thus indicates an association with PAO-D.
**B:** Associations of the PAO (PAO-M and PAO-X *vs.* PAO-D) with the top *vs.* bottom PGS quartiles. A higher OR indicates an association with PAO-D.

**Significance levels:** n.s., *P*>0.05; Nominal, *P*<0.05; Bonferroni, below the Bonferroni-corrected significance threshold corrected for 96 tests (*P*<5.2×10-4).
For detailed results, see Supplementary Table S8.
**Abbreviations:** ADHD, attention deficit/hyperactivity disorder; ASD, autism spectrum disorder; MD, major depression; SZ, schizophrenia; EA, educational attainment; PAO-M, mania/hypomania before depression; PAO-X, mixed episodes or first manic and depressive episode in same year; PAO-D, depression before mania/hypomania.

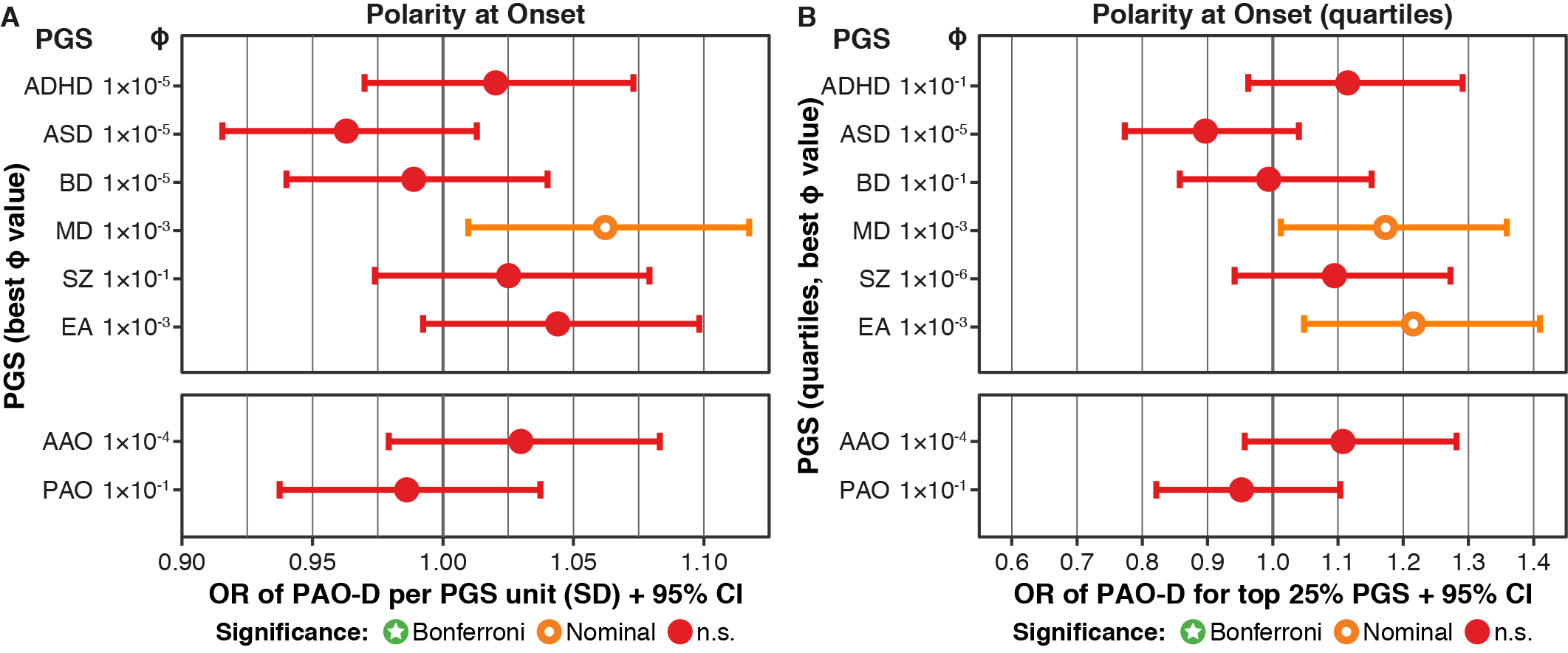

### Supplementary Tables

#### Supplementary Table S1. Overview of the definitions of age at onset (AAO) used by the individual cohorts and their mapping to AAO definition groups in the present manuscript

| Stage | Dataset | Definition of AAO used by the cohort | AAO definition |
| --- | --- | --- | --- |
| Discovery | **wtccc** | Age (years) at first impairment due to an episode of depression, hypomania, mania, or mixed affective episode. | Impairment / help-seeking |
|  | **tgco2** | Age at which the patient first met DSM-IV criteria for a manic, mixed, or major depressive episode. | Diagnostic interview |
|  | **gain** | Age at which proband reported first manic, mixed, or major depressive episode. | Diagnostic interview |
|  | **stp1** | Age at which the patient first met DSM-IV criteria for a manic, mixed, or major depressive episode. | Diagnostic interview |
|  | **gsk1** | Age at which the patient first experienced manic or depressive symptoms, as reported by the patient during the interview | Diagnostic interview |
|  | **usc2** | Age at which psychiatric treatment was first sought or symptoms first began to cause subjective distress or impair functioning, whichever occurred first. | Impairment / help-seeking |
|  | **bonn** | Age at which psychiatric treatment was first sought or symptoms first began to cause subjective distress or impair functioning, whichever occurred first. | Impairment / help-seeking |
|  | **ucl2** | Age at which the patient first received medication to treat a depressive/hypomanic/manic episode | Pharmacotherapy |
|  | **bmg3** | Age at which psychiatric treatment was first sought or symptoms first began to cause subjective distress or impair functioning, whichever occurred first. | Impairment / help-seeking |
|  | ***m&m‘s*** | *PsyCourse (n = 365):* Age at which the patient experienced the first (hypo)manic or depressive episode, based on SCID.  *FOR 2107 (n = 88):* Age at which psychiatric treatment was first sought or when symptoms first began to cause subjective distress or impair functioning, whichever occurred first. | Mixed |
|  | **uclo** | Age at which psychiatric treatment was first sought or when symptoms first began to cause subjective distress or impair functioning, whichever occurred first. | Impairment / help-seeking |
|  | **fran** | Age at which the patient was first reliably diagnosed with a major mood episode (major depression, (hypo)mania, or mixed episode) according to the appropriate section of the DIGS | Diagnostic interview |
|  | **euoR** | *Austria (n = 35):* Age at first subjective symptoms.  *Czech Republic (N = 45):* Age at first illness episode.  *France (n = 46):* Age at which the patient was first reliably diagnosed with a major mood episode (major depression, (hypo)mania or mixed episode) according to the appropriate section of the DIGS.  *Romania (n = 8):* Age at which the patient first met DSM-IV criteria for a manic, mixed, or major depressive episode.  *Spain (n = 73):* Age at which the patient first met DSM-IV criteria for a manic, mixed, or major depressive episode*.*  *Germany (n = 71):* First contact with mental health services because of depressive or manic symptoms.  *Sweden (n = 80):* How old were you when you had your first health care contact for these disorders?  *Switzerland (n = 52):* Age at which the patient first met diagnostic criteria for a manic, mixed, or major depressive episode. | Mixed |
|  | **hal2** | Age at which the patient first met DSM-IV criteria for a manic, mixed, or major depressive episode. | Diagnostic interview |
|  | **ume4** | Age at first ever symptoms of depression, hypomania, or mania on the basis of semi-structured clinical interviews and/or information from clinical records and close relatives. | Diagnostic interview |
|  | **swa2** | How old were you when you had your first health care contact for these disorders? | Impairment / help-seeking |
|  | **bmpo** | Age at which psychiatric treatment was first sought or when symptoms first began to cause subjective distress or impair functioning, whichever occurred first. | Impairment / help-seeking |
|  | **top7** | Age at which the patient first met DSM-IV criteria for a manic, mixed, or major depressive episode. | Diagnostic interview |
|  | **may1** | Age at which the patient first met DSM-IV criteria for a manic, mixed, or major depressive episode. | Diagnostic interview |
|  | **bmsp** | Age at which psychiatric treatment was first sought or when symptoms first began to cause subjective distress or impair functioning, whichever occurred first. | Impairment / help-seeking |
|  | **bmau** | Age at which the patient first met DSM-IV criteria for a manic, mixed, or major depressive episode. | Diagnostic interview |
|  | **edi1** | Recorded as SADS-L items: Age when patient first met criteria for BD | Diagnostic interview |
|  | **rom3** | Age at which the patient first met DSM-IV criteria for a manic, mixed, or major depressive episode. | Diagnostic interview |
|  | **butr** | Age at first impairment caused by symptoms | Impairment / help-seeking |
|  | **euoI** | The first reliably diagnosed (hypo)manic or depressive episode according to RDC criteria, determined by using all available medical records | Diagnostic interview |
|  | **ageu** | How old were you when you had your first health care contact for these disorders? | Impairment / help-seeking |
|  | **mich** | Age at which the patient first met DSM-IV criteria for a manic, mixed, or major depressive episode. | Diagnostic interview |
|  | **naom** | *NIMH (n = 17):* unknown  *Rochester (n = 26):* Age at which the patient first met DSM-IV criteria for a manic, mixed, or major depressive episode.  *Iowa City (n = 13):* Age (self-reported) at which the patient first met the diagnostic criteria for a manic or depressive episode.  *Baltimore (n = 11):* Age (self-reported) at which the patient first met the diagnostic criteria for a manic or depressive episode.  *San Diego (n = 92):* Age at which the patient first met DSM-IV criteria for a manic, mixed, or major depressive episode. | Mixed |
|  | **bmg2** | First contact with mental health services because of depressive or manic symptoms. | Impairment / help-seeking |
|  | **top8** | Age at which the patient first met DSM-IV criteria for a manic, mixed, or major depressive episode. | Diagnostic interview |
|  | **h66x** | *Poland (n = 88):* Age at which psychiatric treatment was first sought OR symptoms first began to cause subjective distress or impair functioning, whichever occurred first.  *Germany (n = 4):* Age at which the patient experienced the first (hypo)manic, mixed or depressive episode, based on SCID. | Mixed |
|  | **auom** | *Adelaide (n = 58):* Age at which the patient first met DSM-IV criteria for a manic, mixed, or major depressive episode.  *Sydney (n = 27):* Age at which the patient first met DSM-IV criteria for a manic, mixed, or major depressive episode. | Diagnostic interview |
|  | **euo2** | Age at which the patient first met the DSM criteria for a mood episode. | Diagnostic interview |
|  | **dub1** | Age at which the patient first met DSM-IV criteria for a manic, mixed, or major depressive episode. | Diagnostic interview |
| Replication | **ukwa1** | Age at which psychiatric treatment was first sought or symptoms first began to cause subjective distress or impair functioning, whichever occurred first. | Impairment / help-seeking |
|  | **dutch** | Age at which the patient first received medication to treat a (hypo)manic, mixed or depressive episode. | Pharmacotherapy |
|  | **jst5** | Unknown | Unknown |
|  | **colo** | Age at which the patient experienced the first (hypo)manic or depressive episode, based on SCID. | Diagnostic interview |
|  | **bmrom** | Age at which the patient first met DSM-IV criteria for a manic, mixed, or major depressive episode. | Diagnostic interview |
|  | **bdtrs** | Age at which psychiatric treatment was first sought | Impairment / help-seeking |

#### Supplementary Table S2. Overview of the genotyping panels and variant counts across the different cohorts used in the primary analyses of age at onset (AAO) and polarity at onset (PAO)

| **Type of GWAS** | **Dataset** | **N** | **Array** | **No. of variants before imputation** | **No. of variants in AAO GWAS** | **No. of variants in AAO meta** | **AAO GWAS λ** | **No. of variants in PAO meta** | **PAO GWAS λ** |
| --- | --- | --- | --- | --- | --- | --- | --- | --- | --- |
| **Discovery** | **wtccc** | 1452 | A5.0 | 432 682 | 8 801 813 | 7 398 963 | 1.017 |  |  |
|  | **tgco2** | 865 | A6.0 | 563 959 | 8 798 153 | 7 563 915 | 1.005 | 7 405 389 | 0.972 |
|  | **gain** | 797 | A6.0 | 677 788 | 8 820 816 | 7 591 369 | 0.998 | 7 429 600 | 0.986 |
|  | **stp1** | 718 | A5.0 | 331 202 | 8 806 379 | 7 498 623 | 0.997 | 7 355 792 | 0.985 |
|  | **gsk1** | 715 | I550 | 528 201 | 8 865 282 | 7 715 207 | 0.999 | 7 546 691 | 0.992 |
|  | **usc2** | 681 | OMEX | 598 185 | 8 985 804 | 7 701 214 | 1.007 |  |  |
|  | **bonn** | 638 | I550 | 499 494 | 8 815 645 | 7 589 780 | 0.984 |  |  |
|  | **ucl2** | 604 | OMEX | 611 804 | 8 818 599 | 7 589 780 | 1.009 | 7 497 908 | 0.998 |
|  | **bmg3** | 455 | I550, I610Q, I660Q | 456 677 | 8 822 557 | 7 589 780 | 1.000 | 7 258 756 | 1.005 |
|  | **m&m’s** | 449 | PsychChip | 244 756 | 8 793 857 | 7 411 897 | 0.995 | 7 208 612 | 0.986 |
|  | **uclo** | 439 | A5.0 | 344 528 | 8 759 835 | 7 351 105 | 1.005 | 7 178 838 | 0.995 |
|  | **fran** | 411 | I650 | 279 572 | 8 788 132 | 7 342 980 | 1.003 |  |  |
|  | **euoR** | 410 | OMEX | 624 675 | 8 922 825 | 7 617 456 | 1.003 |  |  |
|  | **hal2** | 355 | OMEX | 566 260 | 8 848 629 | 7 612 730 | 1.000 | 7 387 863 | 1.002 |
|  | **ume4** | 354 | OMEX | 632 614 | 8 957 109 | 7 612 730 | 1.007 | 7 330 826 | 0.975 |
|  | **swa2** | 344 | A6.0 | 518 940 | 8 851 757 | 7 557 815 | 1.01 |  |  |
|  | **bmpo** | 319 | I317, I660Q | 269 263 | 8 738 828 | 7 557 815 | 1.005 | 7 190 724 | 1.021 |
|  | **top7** | 301 | A6.0 | 667 707 | 8 893 696 | 7 616 405 | 1.005 |  |  |
|  | **may1** | 257 | OMEX | 686 229 | 8 867 638 | 7 618 138 | 1.000 | 7 350 084 | 1.005 |
|  | **bmsp** | 248 | I610Q. I660Q | 329 661 | 9 080 402 | 7 476 861 | 1.003 | 7 303 425 | 1.01 |
|  | **bmau** | 245 | I660Q | 505 360 | 8 788 122 | 7 623 983 | 1.014 | 7 293 919 | 1.004 |
|  | **edi1** | 244 | A5.0 | 344 775 | 8 741 391 | 7 344 672 | 1.008 |  |  |
|  | **rom3** | 226 | OMEX | 587 509 | 8 910 533 | 7 570 414 | 0.997 | 7 337 453 | 1.005 |
|  | **butr** | 204 | OMEX | 656 165 | 8 978 379 | 7 659 546 | 0.993 |  |  |
|  | **euoI** | 191 | OMEX | 622 541 | 8 954 061 | 7 264 673 | 1.016 | 6 974 295 | 1.031 |
|  | **ageu** | 178 | A6.0 | 494 795 | 9 065 654 | 7 635 983 | 0.999 |  |  |
|  | **mich** | 169 | I550 | 509 425 | 8 824 078 | 7 575 360 | 1.009 | 7 328 734 | 1.027 |
|  | **naom** | 159 | OMEX | 624 553 | 8 898 168 | 7 600 346 | 0.996 | 7 090 346 | 1.034 |
|  | **bmg2** | 152 | I01Q | 789 442 | 8 828 658 | 7 561 747 | 1.001 |  |  |
|  | **top8** | 111 | OMEX | 667 049 | 8 903 590 | 7 550 585 | 1.002 |  |  |
|  | **h66x** | 92 | I610Q, I660Q | 412 542 | 8 751 528 | 7 606 230 | 1.018 |  |  |
|  | **auom** | 85 | I660Q | 620 326 | 9 525 561 | 7 722 840 | 1.003 |  |  |
|  | **euo2** | 58 | OMEX | 620 423 | 9 340 134 | 7 722 840 | 0.994 |  |  |
|  | **dub1** | 51 | A6.0 | 660 474 | 8 738 458 | 7 223 862 | 1.007 |  |  |
|  | **Summary** | **12 977** |  |  |  | **7 576 712** | **1.024** | **7 586 624** | **0.97** |
| **Replication** | **ukwa1** | 1156 | PsychChip |  | 7 418 616 | 7 180 534 | 1.001 |  |  |
|  | **dutch** | 468 | OMEX | 302801 | 8 805 961 | 7 595 979 | 0.996 |  |  |
|  | **jst5** | 186 | Il1M |  | 8 865 653 | 7 679 241 | 0.990 |  |  |
|  | **colo** | 176 | GSA | 460009 | 10 560 312 | 8 366 944 | 1.012 |  |  |
|  | **bmrom** | 126 | PsychChip | 536719 | 8 957 237 | 7 543 990 | 1.005 |  |  |
|  | **bdtrs** | 125 | PsychChip | 298475 | 8 800 147 | 7 485 102 | 0.999 |  |  |
|  | **Summary** | **2237** |  |  |  | **7 286 335** | **1.011** |  |  |

GWAS, genome-wide association study; λ, median genomic inflation factor.

#### Supplementary Table S3. Overview of the genome-wide association studies (GWASs) used as training data for single-nucleotide variant weights in the calculation of polygenic scores

| **Phenotype** | **Publication** | **Sample size (cases / controls)** |
| --- | --- | --- |
| Age at onset of bipolar disorder (leave-one-out) | Present study | 12 977 / 0 |
| Attention deficit hyperactivity disorder | Demontis et al. 2019 | 20 183 / 35 191 |
| Autism spectrum disorder | Grove et al. 2019 | 18 381/ 27 969 |
| Bipolar disorder (leave-one-out) | Stahl et al. 2019 | 20 352 / 31 358 |
| Educational attainment | Lee et al. 2018 | 766 345 / 0 |
| Major depressive disorder | Howard et al. 2019 | 170 756 / 329 443 |
| Polarity at onset of bipolar disorder (leave-one-out) | Present Study | 6 773 |
| Schizophrenia | Pardinas et al. 2018 | 40 675 / 64 643 |

#### Supplementary Table S4. Results of separate logistic or linear regression analyses with age at onset (AAO) or polarity at onset (PAO) and sex as independent variables and disease characteristics as dependent variables in the combined analysis of German studies (PsyCourse and FOR2107 cohorts)

| **Variable** |  |  | **AAO** |  |  |  |  | **PAO, including mixed** | |  |  |  |  | **PAO, excluding**  **mixed** | | |  |  |
| --- | --- | --- | --- | --- | --- | --- | --- | --- | --- | --- | --- | --- | --- | --- | --- | --- | --- | --- |
|  | **N** | **OR** | **95% CI** | ***P* value** | **Adj. *P* value** | **N** | **OR** | **95% CI** | ***P* value** | | **Adj. *P* value** | **N** | **OR** | | **95% CI** | ***P* value** | | **Adj. *P* value** |
| **Delusions** | 328 | 0.85 | 0.68-1.07 | 1.67x10^-1^ | 8.35x10^-1^ | 293 | 0.62 | 0.38-1.01 | 5.48x10^-2^ | | 3.29x10^-1^ | 225 | 0.74 | | 0.39-1.39 | 3.45x10^-1^ | | 1.00x10^0^ |
| **Hallucinations** | 336 | 0.80 | 0.62-1.02 | 7.49x10^-2^ | 5.24x10^-1^ | 301 | 0.97 | 0.57-1.65 | 9.21x10^-1^ | | 1.00x10^0^ | 232 | 1.25 | | 0.60-2.58 | 5.52x10^-1^ | | 1.00x10^0^ |
| **Current smoking** | 337 | 0.97 | 0.78-1.20 | 7.72x10^-1^ | 1.00x10^0^ | 302 | 0.84 | 0.53-1.33 | 4.49x10^-1^ | | 1.00x10^0^ | 232 | 0.62 | | 0.33-1.14 | 1.24x10^-1^ | | 6.20x10^-1^ |
| **Suicidal ideation** | 334 | 0.58 | 0.44-0.77 | 1.75x10^-4^ | ***1.58x10^-^***^3^ | 299 | 1.52 | 0.85-2.71 | 1.59x10^-1^ | | 7.95x10^-1^ | 229 | 1.24 | | 0.57-2.68 | 5.92x10^-1^ | | 1.00x10^0^ |
| **Suicide attempt** | 273 | 0.77 | 0.60-1.00 | 5.14x10^-2^ | 4.11x10^-1^ | 256 | 1.80 | 1.06-3.05 | 2.94x10^-2^ | | 2.06x10^-1^ | 186 | 2.37 | | 1.11-5.05 | 2.50x10^-2^ | | 1.50x10^-1^ |
| **Education** | 328 | 0.99 | 0.76-1.28 | 9.12x10^-1^ | 1.00x10^0^ | 294 | 1.31 | 0.76-2.24 | 3.29x10^-1^ | | 1.00x10^0^ | 227 | 1.43 | | 0.70-2.95 | 3.27x10^-1^ | | 1.00x10^0^ |
| **Living together** | 48 | 1.28 | 0.70-2.34 | 4.20x10^-1^ | 1.00x10^0^ | 37 | - | - | - | |  | 37 | - | | - | - | |  |
|  | **N** | **B** | **SE** | ***P* value** | **Adj. *P* value** | **N** | **B** | **SE** | ***P* value** | | **Adj. *P* value** | **N** | **B** | | **SE** | ***P* value** | | **Adj. *P*-value** |
| **Number of manic episodes per year of illness** | 266 | 0.10 | 0.06 | 9.78x10^-2^ | 5.87x10^-1^ | 242 | -0.79 | 0.12 | 3.64x10^-10^ | | ***2.91x10^-9^*** | 187 | -0.51 | | 0.16 | 1.55x10^-3^ | | **1.085x10^-2^** |
| **Number of depressive episodes per year of illness** | 252 | 0.08 | 0.06 | 1.94x10^-1^ | 8.35x10^-1^ | 245 | 0.05 | 0.13 | 7.19x10^-1^ | | 1.00x10^0^ | 189 | 0.52 | | 0.16 | 1.03x10^-3^ | | **8.24x10^-3^** |

The number of manic/depressive episodes was divided by (years of illness)+1. For secondary analyses of the number of episodes not corrected for the years of illness, see the Supplementary Material.

OR, odds ratio; B, unstandardized beta; *P* value, unadjusted *P* value; Adj. *P* value, Bonferroni Holm corrected *P* value; N, total number of cases in model. Significant adjusted *P* values are indicated in bold.

#### Supplementary Table S5. Results of separate logistic or linear regression analyses with age at onset (AAO) or polarity at onset (PAO) and sex as independent and disease characteristics as dependent variables in the Dutch study (ucl2 and Dutch cohorts)

| **Variable** |  |  | **AAO** |  |  |  |  | **PAO, including mixed** | | |  | |  | |  |  | **PAO, excluding mixed** | | | |
| --- | --- | --- | --- | --- | --- | --- | --- | --- | --- | --- | --- | --- | --- | --- | --- | --- | --- | --- | --- | --- |
|  | **N** | **OR** | **95% CI** | ***P* value** | **Adj. *P* value** | **N** | **OR** | | **95% CI** | ***P* value** | | **Adj. *P* value** | | **N** | | **OR** | | **95% CI** | ***P* value** | **Adj. *P* value** |
| **Delusions** | 1284 | 0.67 | 0.59-0.76 | 8.66x10^-10^ | **7.79x10^-9^** | 1005 | 0.62 | | 0.48-0.82 | 7.43x10^-4^ | | **5.20x10^-3^** | | 545 | | 0.19 | | 0.09-0.39 | 5x10^-6^ | **4.5x10^-5^** |
| **Hallucinations** | 1258 | 0.83 | 0.74-0.93 | 1.83x10^-3^ | **7.32x10^-3^** | 989 | 0.92 | | 0.71-1.18 | 5.08x10^-1^ | | 1.00x10^0^ | | 541 | | 0.70 | | 0.45-1.09 | 1.16x10^-1^ | 3.48x10^-1^ |
| **Current smoking** | 1257 | 0.99 | 0.88-1.11 | 8.38x10^-1^ | 8.38x10^-1^ | 980 | 1.23 | | 0.94-1.60 | 1.24x10^-1^ | | 4.96x10^-1^ | | 531 | | 1.03 | | 0.65-1.64 | 9.01x10^-1^ | 9.24x10^-1^ |
| **Suicidal ideation** | 1184 | 0.83 | 0.74-0.94 | 2.60x10^-3^ | **7.80x10^-3^** | 981 | 1.71 | | 1.32-2.23 | 5.6x10^-5^ | | **4.48x10^-4^** | | 545 | | 2.09 | | 1.34-3.27 | 1.18x10^-3^ | **8.26x10^-3^** |
| **Suicide attempt** | 1264 | 0.78 | 0.68-0.89 | 2.03x10^-4^ | **1.42x10^-3^** | 1006 | 1.52 | | 1.15-2.01 | 2.94x10^-3^ | | **1.76x10^-2^** | | 550 | | 1.94 | | 1.14-3.31 | 1.427x10^-2^ | 8.58x10^-2^ |
| **Education** | 1308 | 1.21 | 1.08-1.35 | 9.51x10^-4^ | **4.76x10^-3^** | 1025 | 1.02 | | 0.79-1.30 | 8.88x10^-1^ | | 1.00x10^0^ | | 557 | | 0.67 | | 0.43-1.04 | 7.23x10^-2^ | 2.89x10^-1^ |
| **Living together** | 1309 | 1.28 | 1.15-1.44 | 1.40x10^-5^ | **1.12x10^-4^** | 1025 | 0.86 | | 0.67-1.10 | 2.29x10^-1^ | | 6.87x10^-1^ | | 557 | | 1.18 | | 0.76-1.82 | 4.62x10^-1^ | 9.24x10^-1^ |
|  | **N** | **B** | **SE** | ***P* value** | **Adj. *P* value** | **N** | **B** | | **SE** | ***P* value** | | **Adj. *P* value** | | **N** | | **B** | | **SE** | ***P* value** | **Adj. *P* value** |
| **Number of manic episodes per year of illness** | 1171 | 0.11 | 0.03 | 3.17x10^-4^ | **1.90x10^-3^** | 916 | -0.31 | | 0.07 | 4x10^-6^ | | **3.6x10^-5^** | | 498 | | -0.25 | | 0.12 | 3.03x10^-2^ | 1.515x10^-1^ |
| **Number of depressive episodes per year of illness** | 981 | 0.06 | 0.03 | 5.04x10^-2^ | 1.01x10^-1^ | 808 | 0.14 | | 0.07 | 3.91x10^-2^ | | 1.96x10^-1^ | | 445 | | 0.35 | | 0.11 | 1.01x10^-3^ | **8.08x10^-3^** |

The number of manic/depressive episodes was divided by (years of illness)+1. For secondary analyses of the number of episodes not corrected for the years of illness, see the Supplementary Material.

OR, odds ratio; B, unstandardized beta; *P* value, unadjusted *P* value; Adj. *P* value, Bonferroni Holm corrected *P* value; N, total number of cases in model. Significant adjusted *P* values are indicated in bold.

#### Supplementary Table S6: Differences in age at onset (AAO) between subgroups

Non-parametric pairwise Mann-Whitney U tests (median, *χ^2^* statistic (chi^2), *P* value) and linear regression model (beta, SE, *P* value) on the untransformed age at onset.

For these analyses, the default definition of AAO was the diagnostic interview; the default continent, Europe; the default subtype, BD-I; and the default sex, male.

Separate Mann-Whitney U tests were used to assess continent, definition, subtype, and sex. A single, multivariable model containing all listed variables was used for the linear regression. Thus, the coefficients from the linear regression model are corrected for the other variables displayed, while the Mann-Whitney U test results are univariate.

|  | **Mann-Whitney U tests** | | | **Linear regression** | | |
| --- | --- | --- | --- | --- | --- | --- |
| **Variable** | **Median** | **Chi^2** | ***P* value** | **Beta** | **SE** | ***P* value** |
| **Continent: Europe** | 24 |  |  |  |  |  |
| **Continent: North America** | 18 | 1202.3 | **1.97E-263** | -4.70 | 0.22 | **5.12E-96** |
| **Continent: Australia** | 19.5 | 47.69 | **4.97E-12** | -2.18 | 0.58 | **1.54E-04** |
| **Definition: Diagnostic interview** | 19 |  |  |  |  |  |
| **Definition: Impairment/help-seeking** | 23 | 517.64 | **1.38E-114** | 1.44 | 0.22 | **1.32E-10** |
| **Definition: Pharmacotherapy** | 30 | 490.29 | **1.23E-108** | 6.73 | 0.45 | **3.40E-50** |
| **Definition: Mixed** | 23 | 143.95 | **3.63E-33** | -1.85 | 1.39 | 1.84E-01 |
| **Subtype: BD-I** | 21 |  |  |  |  |  |
| **Subtype: BD-II** | 22 | 17.13 | **3.48E-05** | 0.86 | 0.28 | **2.43E-03** |
| **Subtype: BDNOS** | 20 | 0.04 | 0.835 | 0.97 | 1.05 | 3.55E-01 |
| **Sex: Male** | 22 |  |  |  |  |  |
| **Sex: Female** | 21 | 23.79 | **1.07E-06** | -0.86 | 0.18 | **1.11E-06** |

BDNOS, bipolar disorder not otherwise specified

#### Supplementary Table S7: Genome-wide association study on age at onset (AAO) in bipolar disorder

Genome-wide significant locus at rs1610275 on chromosome 16 for AAO in the primary and replication analyses.

|  | **Allele Frequency** | **INFO Score** | **Beta** | **SE** | ***P*** | **N** |
| --- | --- | --- | --- | --- | --- | --- |
| **Discovery** | 0.319 (G) | 0.969 | 0.075 | 0.0135 | 3.388E-08 | 12977 |
| **Replication** | 0.321 (G) | 0.982 | -0.042 | 0.0329 | 0.1929 | 2237 |

#### Supplementary Table S8: Results of PGS analyses

Please see the separate Excel file.

Significance levels: Bonferroni, significant after correction for 96 tests, *P* < 5.2×10^-4^; Nominal, *P* < 0.05; n.s., not significant. *P* 1-sided: one-sided *P* value, based on the hypothesis that all polygenic scores (PGSs) except the PGS for age at onset (AAO) show a negative association with AAO. R^2 complete: *R^2^* complete model; R^2 null: *R^2^* null model (without PGS); R^2 PGS: *R^2^* explained by the PGS; N. R^2: Nagelkerke’s pseudo-*R^2^*. N: sample size; I^2^/Q: Measures of meta-analysis heterogeneity.

#### Supplementary Table S9. Overview of characteristics for phenotypic analyses

|  | **PsyCourse** | |  | **FOR2107** |  | **Dutch BP** |  | **Outcome variable** |
| --- | --- | --- | --- | --- | --- | --- | --- | --- |
|  | **Instrument** | **Items** | | **Instrument** | **Items** | **Instrument** | **Items** |  |
| **Delusions** | SCID-I(4) | Delusion of reference  Persecutory delusion  Delusions of grandiosity  Somatic delusion  Other delusion  Delusion of control  Thought withdrawal  Religious delusion  Delusion of guilt  Delusion of jealousy  Erotomanic delusion  Cotard delusion  Delusion of poverty | | SCID-I | Delusion of reference  Persecutory delusion  Delusions of grandiosity  Somatic delusion  Other delusion  Ego disturbance  Thought broadcast | SCID-I* | Delusion of reference  Persecutory delusion  Delusion of grandiosity  Somatic delusion  Other delusion  Religious delusion  Delusion of guilt or sin  Delusion of jealousy  Erotomanic delusion  Delusion of being controlled  Thought insertion  Thought withdrawal  Thought broadcasting  Bizarre delusion | Delusions lifetime; dichotomous  [No, has never experienced any of delusions asked about = 0; yes, has experienced at least one of the delusions asked about = 1] |
| **Hallucinations** | SCID-I | Auditory hallucinations  Visual hallucinations  Olfactory hallucinations  Gustatory hallucinations  Tactile hallucinations/ | | SCID-I | Auditory hallucinations  Visual hallucinations  Tactile hallucinations  Other hallucinations | SCID-I | Auditory hallucinations  Visual hallucinations  Tactile hallucinations  Other hallucinations | Lifetime hallucinations;  dichotomous [No = 0; Yes = 1] |
| **Current smoking** | Structured interview | 1 item: Have you ever smoked cigarettes, cigars, pipe, or other tobacco products?  [1. Never smoked (or < 100 cigarettes during lifetime); 2. Yes, current smoker; 3. Former smoker (quit smoking more than 3 months ago)] | | Fagerström (self-report) | 1 item: Current smoker? | Fagerström (self-report)(5) | 1 item: Do you smoke currently? | Current smoking;  dichotomous [No = 0; Yes = 1] |
| **Suicidal ideation** | SCID-I | Suicidal ideation, lifetime | | OPCRIT | Suicidal ideation, lifetime | SCID-I | Suicidal ideation during depressive episode | Suicidal ideation; dichotomous [No = 0; Yes = 1] |
| **Suicide attempt** | SCID-I | Suicide attempt, lifetime  [No = 1; interrupted attempt = 2; yes = 3] | | - |  | Combination of items SCID-I and Comprehensive Assessment of Symptoms and History (CASH)(6) | SCID-I suicide attempt during depressive episode:  CASH: Suicide attempt, lifetime | Lifetime suicide attempt; dichotomous [No = 0; Yes = 1] |
| **Education** | Interview  *Education (ordinal [0,1,2,3,4,5,6], v1_ed_status):*  This scale was newly created by merging the original items. **    * High school-level education (categorical [0,1,2,3], v1_school).  no information/missing = NA;  no graduation = 0; high school completed after grade 9 = 1;  high school completed after grade 10 OR polytechnic high school = 2;  technical high school OR European general higher education entrance qualification = 3;  still in school/other type of school diploma = 999;  * Professional education (categorical [0,1,2,3]  missing or no information = NA;  no professional education/vocational training in a company but no apprenticeship/ vocational training program/in professional education = 0; apprenticeship = 1; vocational training in a company /vocational and school-based training = 2; degree from a university or university of applied sciences = 3; other professional degree = 999,  Important: more than 1 answer was possible, because people may have several professional degrees. |  | | Interview  Highest completed educational level  No school diploma = 1;  elementary school = 2;  diploma from high school completed after grade 9 = 3; high school completed after grade 10 = 4;  technical diploma = 5;  high school diploma = 6;  apprenticeship = 7;  master craftsman = 8; Bachelors = 9;  Masters = 10 |  | Self-report questionnaire  [Low education = 1; intermediate secondary education = 2; intermediate professional education = 3; high preparatory vocational/pre-university = 4; Bachelor = 5; Master or PhD degree = 6] |  | Educational attainment, dichotomous  [Lower educational attainment ((PsyCourse: 0, 1, 2, 3, 4, 5; FOR: 1, 2, 3, 4, 5, 6, 7, 8; Dutch BP: 1, 2, 3, 4)= 0); Higher educational attainment (PsyCourse: 6; FOR: 9, 10, Dutch BP: 5, 6)= 1] |
| **Living together** | - |  | | SCID-I | Current living situation  [Living alone = 1; living with partner = 2; living with husband/wife = 3; living with parents/relatives = 4; living in a community = 5; living in a treatment facility = 6; other = 7]  5, 6, and 7 are recoded as missing because in these cases it is unclear whether someone is living with a partner | SCID-I | Current marital status  [Married or living together = 1; widowed = 2; divorced = 3; divorce from bed and board = 4; never married = 5] | Living together, dichotomous [Not living together with a partner (living alone, living with parents/relatives divorced, divorced from bed and board, never married) = 0; living together with a partner or widowed (living with a partner, living with husband/wife, married or living together, widowed) = 1] |
| **Number of manic** | SCID-I | Total number of manic episodes | | SCID-I | Total number of manic episodes | SCID-I | Total number of manic episodes | Rank normalized variable number of episodes; continuous  number of episodes/(years of illness + 1) |
| **Number of depressive episodes** | SCID-I | Total number of depressive episodes | | SCID-I | Total number of depressive episodes | SCID-I | Total number of depressive episodes | Rank normalized variable number of episodes; continuous  number of episodes/(years of illness + 1) |

SCID-I, Structured Clinical Interview for DSM-IV; CASH, Comprehensive Assessment of Symptoms and History

*Questionable is coded as absent. However, if screener questions are coded questionable, but specifier items are coded as present, then Delusions lifetime and Hallucinations lifetime are coded as present.

** School and university/professional education were assessed separately in the interview. To combine school and university/ professional education, we transformed both scales to values that could be added together to form an “educational attainment” variable. High school-level education was transformed into an ordinal scale from 0 to 3 (people still in high school at the time of the interview were assigned “NA”). University/professional education was also transformed into an ordinal scale from 0 to 3. These 2 scales were added together to give an ordinal educational status scale, which ranged from 0 to 6.

### Authors of the Bipolar Disorder Working Group of the Psychiatric Genomics Consortium

Eli A Stahl 1,2,3†

Gerome Breen 4,5†

Andreas J Forstner 6,7,8,9,10†

Andrew McQuillin 11†

Stephan Ripke 12,13,14†

Vassily Trubetskoy 13

Manuel Mattheisen 15,16,17,18,19

Yunpeng Wang 20,21

Jonathan R I Coleman 4,5

Héléna A Gaspar 4,5

Christiaan A de Leeuw 22

Stacy Steinberg 23

Jennifer M Whitehead Pavlides 24

Maciej Trzaskowski 25

Tune H Pers 3,26

Peter A Holmans 27

Liam Abbott 12

Esben Agerbo 19,28,29

Huda Akil 30

Diego Albani 31

Ney Alliey-Rodriguez 32

Thomas D Als 15,16,19

Adebayo Anjorin 33

Verneri Antilla 14

Swapnil Awasthi 13

Judith A Badner 34

Marie Bækvad-Hansen 19,35

Jack D Barchas 36

Nicholas Bass 11

Michael Bauer 37

Richard Belliveau 12

Sarah E Bergen 38

Carsten Bøcker Pedersen 19,28,29

Erlend Bøen 39

Marco Boks 40

James Boocock 41

Monika Budde 42

William Bunney 43

Margit Burmeister 44

Jonas Bybjerg-Grauholm 19,35

William Byerley 45

Miquel Casas 46,47,48,49

Felecia Cerrato 12

Pablo Cervantes 50

Kimberly Chambert 12

Alexander W Charney 2

Danfeng Chen 12

Claire Churchhouse 12,14

Toni-Kim Clarke 51

William Coryell 52

David W Craig 53

Cristiana Cruceanu 50,54

Piotr M Czerski 55

Anders M Dale 56,57,58,59

Simone de Jong 4,5

Franziska Degenhardt 8,9

Jurgen Del-Favero 60

J Raymond DePaulo 61

Srdjan Djurovic 62,63

Amanda L Dobbyn 1,2

Ashley Dumont 12

Torbjørn Elvsåshagen 64,65

Valentina Escott-Price 27

Chun Chieh Fan 59

Sascha B Fischer 6,10

Matthew Flickinger 66

Tatiana M Foroud 67

Liz Forty 27

Josef Frank 68

Christine Fraser 27

Nelson B Freimer 69

Louise Frisén 70,71,72

Katrin Gade 42,73

Diane Gage 12

Julie Garnham 74

Claudia Giambartolomei 41

Marianne Giørtz Pedersen 19,28,29

Jaqueline Goldstein 12

Scott D Gordon 75

Katherine Gordon-Smith 76

Elaine K Green 77

Melissa J Green 78

Tiffany A Greenwood 58

Jakob Grove 15,16,19,79

Weihua Guan 80

José Guzman Parra 81

Marian L Hamshere 27

Martin Hautzinger 82

Urs Heilbronner 42

Stefan Herms 6,8,9,10

Maria Hipolito 83

Per Hoffmann 6,8,9,10

Dominic Holland 56,84

Laura Huckins 1,2

Stéphane Jamain 85,86

Jessica S Johnson 1,2

Anders Juréus 38

Radhika Kandaswamy 4

Robert Karlsson 38

James L Kennedy 87,88,89,90

Sarah Kittel-Schneider 91

James A Knowles 92,93

Manolis Kogevinas 94

Anna C Koller 8,9

Ralph Kupka 95,96,97

Catharina Lavebratt 70

Jacob Lawrence 98

William B Lawson 83

Markus Leber 99

Phil H Lee 12,14,100

Shawn E Levy 101

Jun Z Li 102

Chunyu Liu 103

Susanne Lucae 104

Anna Maaser 8,9

Donald J MacIntyre 105,106

Pamela B Mahon 61,107

Wolfgang Maier 108

Lina Martinsson 71

Steve McCarroll 12,109

Peter McGuffin 4

Melvin G McInnis 110

James D McKay 111

Helena Medeiros 93

Sarah E Medland 75

Fan Meng 30,110

Lili Milani 112

Grant W Montgomery 25

Derek W Morris 113,114

Thomas W Mühleisen 6,115

Niamh Mullins 4

Hoang Nguyen 1,2

Caroline M Nievergelt 58,116

Annelie Nordin Adolfsson 117

Evaristus A Nwulia 83

Claire O'Donovan 74

Loes M Olde Loohuis 69

Anil P S Ori 69

Lilijana Oruc 118

Urban Ösby 119

Roy H Perlis 120,121

Amy Perry 76

Andrea Pfennig 37

James B Potash 61

Shaun M Purcell 2,107

Eline J Regeer 122

Andreas Reif 91

Céline S Reinbold 6,10

John P Rice 123

Alexander L Richards 27

Fabio Rivas 81

Margarita Rivera 4,124

Panos Roussos 1,2,125

Douglas M Ruderfer 126

Euijung Ryu 127

Cristina Sánchez-Mora 46,47,49

Alan F Schatzberg 128

William A Scheftner 129

Nicholas J Schork 130

Cynthia Shannon Weickert 78,131

Tatyana Shehktman 58

Paul D Shilling 58

Engilbert Sigurdsson 132

Claire Slaney 74

Olav B Smeland 56,133,134

Janet L Sobell 135

Christine Søholm Hansen 19,35

Anne T Spijker 136

David St Clair 137

Michael Steffens 138

John S Strauss 89,139

Fabian Streit 68

Jana Strohmaier 68

Szabolcs Szelinger 140

Robert C Thompson 110

Thorgeir E Thorgeirsson 23

Jens Treutlein 68

Helmut Vedder 141

Weiqing Wang 1,2

Stanley J Watson 110

Thomas W Weickert 78,131

Stephanie H Witt 68

Simon Xi 142

Wei Xu 143,144

Allan H Young 145

Peter Zandi 146

Peng Zhang 147

Sebastian Zollner 110

Rolf Adolfsson 117

Ingrid Agartz 17,39,148

Martin Alda 74,149

Lena Backlund 71

Bernhard T Baune 150

Frank Bellivier 151,152,153,154

Wade H Berrettini 155

Joanna M Biernacka 127

Douglas H R Blackwood 51

Michael Boehnke 66

Anders D Børglum 15,16,19

Aiden Corvin 114

Nicholas Craddock 27

Mark J Daly 12,14

Udo Dannlowski 156

Tõnu Esko 3,109,112,157

Bruno Etain 151,153,154,158

Mark Frye 159

Janice M Fullerton 131,160

Elliot S Gershon 32,161

Michael Gill 114

Fernando Goes 61

Maria Grigoroiu-Serbanescu 162

Joanna Hauser 55

David M Hougaard 19,35

Christina M Hultman 38

Ian Jones 27

Lisa A Jones 76

René S Kahn 2,40

George Kirov 27

Mikael Landén 38,163

Marion Leboyer 86,151,164

Cathryn M Lewis 4,5,165

Qingqin S Li 166

Jolanta Lissowska 167

Nicholas G Martin 75,168

Fermin Mayoral 81

Susan L McElroy 169

Andrew M McIntosh 51,170

Francis J McMahon 171

Ingrid Melle 172,173

Andres Metspalu 112,174

Philip B Mitchell 78

Gunnar Morken 175,176

Ole Mors 19,177

Preben Bo Mortensen 15,19,28,29

Bertram Müller-Myhsok 54,178,179

Richard M Myers 101

Benjamin M Neale 3,12,14

Vishwajit Nimgaonkar 180

Merete Nordentoft 19,181

Markus M Nöthen 8,9

Michael C O'Donovan 27

Ketil J Oedegaard 182,183

Michael J Owen 27

Sara A Paciga 184

Carlos Pato 93,185

Michele T Pato 93

Danielle Posthuma 22,186

Josep Antoni Ramos-Quiroga 46,47,48,49

Marta Ribasés 46,47,49

Marcella Rietschel 68

Guy A Rouleau 187,188

Martin Schalling 70

Peter R Schofield 131,160

Thomas G Schulze 42,61,68,73,171

Alessandro Serretti 189

Jordan W Smoller 12,190,191

Hreinn Stefansson 23

Kari Stefansson 23,192

Eystein Stordal 193,194

Patrick F Sullivan 38,195,196

Gustavo Turecki 197

Arne E Vaaler 198

Eduard Vieta 199

John B Vincent 139

Thomas Werge 19,200,201

John I Nurnberger 202

Naomi R Wray 24,25

Arianna Di Florio 27,196

Howard J Edenberg 203

Sven Cichon 6,8,10,115

Roel A Ophoff 40,41,69

Laura J Scott 66

Ole A Andreassen 133,134

John Kelsoe 58*

Pamela Sklar 1,2*^

† Equal contribution, * Co-last authors

^ deceased.

1. Department of Genetics and Genomic Sciences, Icahn School of Medicine at Mount Sinai, New York, NY, US
2. Department of Psychiatry, Icahn School of Medicine at Mount Sinai, New York, NY, US
3. Medical and Population Genetics, Broad Institute, Cambridge, MA, US
4. MRC Social, Genetic and Developmental Psychiatry Centre, King's College London, London, GB
5. NIHR BRC for Mental Health, King's College London, London, GB
6. Human Genomics Research Group, Department of Biomedicine, University of Basel, Basel, CH
7. Department of Psychiatry (UPK), University of Basel, Basel, CH
8. Institute of Human Genetics, University of Bonn, School of Medicine & University Hospital Bonn, Bonn, DE
9. Department of Genomics, Life&Brain Center, University of Bonn, Bonn, DE
10. Institute of Medical Genetics and Pathology, University Hospital Basel, Basel, CH
11. Division of Psychiatry, University College London, London, GB
12. Stanley Center for Psychiatric Research, Broad Institute, Cambridge, MA, US
13. Department of Psychiatry and Psychotherapy, Charité - Universitätsmedizin, Berlin, DE
14. Analytic and Translational Genetics Unit, Massachusetts General Hospital, Boston, MA, US
15. iSEQ, Center for Integrative Sequencing, Aarhus University, Aarhus, DK
16. Department of Biomedicine - Human Genetics, Aarhus University, Aarhus, DK
17. Department of Clinical Neuroscience, Centre for Psychiatry Research, Karolinska Institutet, Stockholm, SE
18. Department of Psychiatry, Psychosomatics and Psychotherapy, Center of Mental Health, University Hospital Würzburg, Würzburg, DE
19. iPSYCH, The Lundbeck Foundation Initiative for Integrative Psychiatric Research, DK
20. Institute of Biological Psychiatry, Mental Health Centre Sct. Hans, Copenhagen, DK
21. Institute of Clinical Medicine, University of Oslo, Oslo, NO
22. Department of Complex Trait Genetics, Center for Neurogenomics and Cognitive Research, Amsterdam Neuroscience, Vrije Universiteit Amsterdam, Amsterdam, NL
23. deCODE Genetics / Amgen, Reykjavik, IS
24. Queensland Brain Institute, The University of Queensland, Brisbane, QLD, AU
25. Institute for Molecular Bioscience, The University of Queensland, Brisbane, QLD, AU
26. Division of Endocrinology and Center for Basic and Translational Obesity Research, Boston Children’s Hospital, Boston, MA, US
27. Medical Research Council Centre for Neuropsychiatric Genetics and Genomics, Division of Psychological Medicine and Clinical Neurosciences, Cardiff University, Cardiff, GB
28. National Centre for Register-Based Research, Aarhus University, Aarhus, DK
29. Centre for Integrated Register-based Research, Aarhus University, Aarhus, DK
30. Molecular & Behavioral Neuroscience Institute, University of Michigan, Ann Arbor, MI, US
31. NEUROSCIENCE, Istituto Di Ricerche Farmacologiche Mario Negri, Milano, IT
32. Department of Psychiatry and Behavioral Neuroscience, University of Chicago, Chicago, IL, US
33. Psychiatry, Berkshire Healthcare NHS Foundation Trust, Bracknell, GB
34. Psychiatry, Rush University Medical Center, Chicago, IL, US
35. Center for Neonatal Screening, Department for Congenital Disorders, Statens Serum Institut, Copenhagen, DK
36. Department of Psychiatry, Weill Cornell Medical College, New York, NY, US
37. Department of Psychiatry and Psychotherapy, University Hospital Carl Gustav Carus, Technische Universität Dresden, Dresden, DE
38. Department of Medical Epidemiology and Biostatistics, Karolinska Institutet, Stockholm, SE
39. Department of Psychiatric Research, Diakonhjemmet Hospital, Oslo, NO
40. Psychiatry, UMC Utrecht Hersencentrum Rudolf Magnus, Utrecht, NL
41. Human Genetics, University of California Los Angeles, Los Angeles, CA, US
42. Institute of Psychiatric Phenomics and Genomics (IPPG), University Hospital, LMU Munich, Munich, DE
43. Department of Psychiatry and Human Behavior, University of California, Irvine, Irvine, CA, US
44. Molecular & Behavioral Neuroscience Institute and Department of Computational Medicine & Bioinformatics, University of Michigan, Ann Arbor, MI, US
45. Psychiatry, University of California San Francisco, San Francisco, CA, US
46. Instituto de Salud Carlos III, Biomedical Network Research Centre on Mental Health (CIBERSAM), Madrid, ES
47. Department of Psychiatry, Hospital Universitari Vall d´Hebron, Barcelona, ES
48. Department of Psychiatry and Forensic Medicine, Universitat Autònoma de Barcelona, Barcelona, ES
49. Psychiatric Genetics Unit, Group of Psychiatry Mental Health and Addictions, Vall d´Hebron Research Institut (VHIR), Universitat Autònoma de Barcelona, Barcelona, ES
50. Department of Psychiatry, Mood Disorders Program, McGill University Health Center, Montreal, QC, CA
51. Division of Psychiatry, University of Edinburgh, Edinburgh, GB
52. University of Iowa Hospitals and Clinics, Iowa City, IA, US
53. Translational Genomics, USC, Phoenix, AZ, US
54. Department of Translational Research in Psychiatry, Max Planck Institute of Psychiatry, Munich, DE
55. Department of Psychiatry, Laboratory of Psychiatric Genetics, Poznan University of Medical Sciences, Poznan, PL
56. Department of Neurosciences, University of California San Diego, La Jolla, CA, US
57. Department of Radiology, University of California San Diego, La Jolla, CA, US
58. Department of Psychiatry, University of California San Diego, La Jolla, CA, US
59. Department of Cognitive Science, University of California San Diego, La Jolla, CA, US
60. Applied Molecular Genomics Unit, VIB Department of Molecular Genetics, University of Antwerp, Antwerp, Belgium
61. Department of Psychiatry and Behavioral Sciences, Johns Hopkins University School of Medicine, Baltimore, MD, US
62. Department of Medical Genetics, Oslo University Hospital Ullevål, Oslo, NO
63. NORMENT, KG Jebsen Centre for Psychosis Research, Department of Clinical Science, University of Bergen, Bergen, NO
64. Department of Neurology, Oslo University Hospital, Oslo, NO
65. NORMENT, KG Jebsen Centre for Psychosis Research, Oslo University Hospital, Oslo, NO
66. Center for Statistical Genetics and Department of Biostatistics, University of Michigan, Ann Arbor, MI, US
67. Department of Medical & Molecular Genetics, Indiana University, Indianapolis, IN, US
68. Department of Genetic Epidemiology in Psychiatry, Central Institute of Mental Health, Medical Faculty Mannheim, Heidelberg University, Mannheim, DE
69. Center for Neurobehavioral Genetics, University of California Los Angeles, Los Angeles, CA, US
70. Department of Molecular Medicine and Surgery, Karolinska Institutet and Center for Molecular Medicine, Karolinska University Hospital, Stockholm, SE
71. Department of Clinical Neuroscience, Karolinska Institutet and Center for Molecular Medicine, Karolinska University Hospital, Stockholm, SE
72. Child and Adolescent Psychiatry Research Center, Stockholm, SE
73. Department of Psychiatry and Psychotherapy, University Medical Center Göttingen, Göttingen, DE
74. Department of Psychiatry, Dalhousie University, Halifax, NS, CA
75. Genetics and Computational Biology, QIMR Berghofer Medical Research Institute, Brisbane, QLD, AU
76. Department of Psychological Medicine, University of Worcester, Worcester, GB
77. School of Biomedical and Healthcare Sciences, Plymouth University Peninsula Schools of Medicine and Dentistry, Plymouth, GB
78. School of Psychiatry, University of New South Wales, Sydney, NSW, AU
79. Bioinformatics Research Centre, Aarhus University, Aarhus, DK
80. Biostatistics, University of Minnesota System, Minneapolis, MN, US
81. Mental Health Department, University Regional Hospital, Biomedicine Institute (IBIMA), Málaga, ES
82. Department of Psychology, Eberhard Karls Universität Tübingen, Tubingen, DE
83. Department of Psychiatry and Behavioral Sciences, Howard University Hospital, Washington, DC, US
84. Center for Multimodal Imaging and Genetics, University of California San Diego, La Jolla, CA, US
85. Psychiatrie Translationnelle, Inserm U955, Créteil, FR
86. Faculté de Médecine, Université Paris Est, Créteil, FR
87. Campbell Family Mental Health Research Institute, Centre for Addiction and Mental Health, Toronto, ON, CA
88. Neurogenetics Section, Centre for Addiction and Mental Health, Toronto, ON, CA
89. Department of Psychiatry, University of Toronto, Toronto, ON, CA
90. Institute of Medical Sciences, University of Toronto, Toronto, ON, CA
91. Department of Psychiatry, Psychosomatic Medicine and Psychotherapy, University Hospital Frankfurt, Frankfurt am Main, DE
92. Cell Biology, SUNY Downstate Medical Center College of Medicine, Brooklyn, NY, US
93. Institute for Genomic Health, SUNY Downstate Medical Center College of Medicine, Brooklyn, NY, US
94. ISGlobal, Barcelona, ES
95. Psychiatry, Altrecht, Utrecht, NL
96. Psychiatry, GGZ inGeest, Amsterdam, NL
97. Psychiatry, VU medisch centrum, Amsterdam, NL
98. Psychiatry, North East London NHS Foundation Trust, Ilford, GB
99. Clinic for Psychiatry and Psychotherapy, University Hospital Cologne, Cologne, DE
100. Psychiatric and Neurodevelopmental Genetics Unit, Massachusetts General Hospital, Boston, MA, US
101. HudsonAlpha Institute for Biotechnology, Huntsville, AL, US
102. Department of Human Genetics, University of Michigan, Ann Arbor, MI, US
103. Psychiatry, University of Illinois at Chicago College of Medicine, Chicago, IL, US
104. Max Planck Institute of Psychiatry, Munich, DE
105. Mental Health, NHS 24, Glasgow, GB
106. Division of Psychiatry, Centre for Clinical Brain Sciences, University of Edinburgh, Edinburgh, GB
107. Psychiatry, Brigham and Women's Hospital, Boston, MA, US
108. Department of Psychiatry and Psychotherapy, University of Bonn, Bonn, DE
109. Department of Genetics, Harvard Medical School, Boston, MA, US
110. Department of Psychiatry, University of Michigan, Ann Arbor, MI, US
111. Genetic Cancer Susceptibility Group, International Agency for Research on Cancer, Lyon, FR
112. Estonian Genome Center, University of Tartu, Tartu, EE
113. Discipline of Biochemistry, Neuroimaging and Cognitive Genomics (NICOG) Centre, National University of Ireland, Galway, Galway, IE
114. Neuropsychiatric Genetics Research Group, Dept of Psychiatry and Trinity Translational Medicine Institute, Trinity College Dublin, Dublin, IE
115. Institute of Neuroscience and Medicine (INM-1), Research Centre Jülich, Jülich, DE
116. Research/Psychiatry, Veterans Affairs San Diego Healthcare System, San Diego, CA, US
117. Department of Clinical Sciences, Psychiatry, Umeå University Medical Faculty, Umeå, SE
118. Department of Clinical Psychiatry, Psychiatry Clinic, Clinical Center University of Sarajevo, Sarajevo, BA
119. Department of Neurobiology, Care sciences, and Society, Karolinska Institutet and Center for Molecular Medicine, Karolinska University Hospital, Stockholm, SE
120. Psychiatry, Harvard Medical School, Boston, MA, US
121. Division of Clinical Research, Massachusetts General Hospital, Boston, MA, US
122. Outpatient Clinic for Bipolar Disorder, Altrecht, Utrecht, NL
123. Department of Psychiatry, Washington University in Saint Louis, Saint Louis, MO, US
124. Department of Biochemistry and Molecular Biology II, Institute of Neurosciences, Center for Biomedical Research, University of Granada, Granada, ES
125. Department of Neuroscience, Icahn School of Medicine at Mount Sinai, New York, NY, US
126. Medicine, Psychiatry, Biomedical Informatics, Vanderbilt University Medical Center, Nashville, TN, US
127. Department of Health Sciences Research, Mayo Clinic, Rochester, MN, US
128. Psychiatry and Behavioral Sciences, Stanford University School of Medicine, Stanford, CA, US
129. Rush University Medical Center, Chicago, IL, US
130. Scripps Translational Science Institute, La Jolla, CA, US
131. Neuroscience Research Australia, Sydney, NSW, AU
132. Faculty of Medicine, Department of Psychiatry, School of Health Sciences, University of Iceland, Reykjavik, IS
133. Div Mental Health and Addiction, Oslo University Hospital, Oslo, NO
134. NORMENT, University of Oslo, Oslo, NO
135. Psychiatry and the Behavioral Sciences, University of Southern California, Los Angeles, CA, US
136. Mood Disorders, PsyQ, Rotterdam, NL
137. Institute for Medical Sciences, University of Aberdeen, Aberdeen, UK
138. Research Division, Federal Institute for Drugs and Medical Devices (BfArM), Bonn, DE
139. Centre for Addiction and Mental Health, Toronto, ON, CA
140. Neurogenomics, TGen, Los Angeles, AZ, US
141. Psychiatry, Psychiatrisches Zentrum Nordbaden, Wiesloch, DE
142. Computational Sciences Center of Emphasis, Pfizer Global Research and Development, Cambridge, MA, US
143. Department of Biostatistics, Princess Margaret Cancer Centre, Toronto, ON, CA
144. Dalla Lana School of Public Health, University of Toronto, Toronto, ON, CA
145. Psychological Medicine, Institute of Psychiatry, Psychology & Neuroscience, King's College London, London, GB
146. Department of Mental Health, Johns Hopkins University Bloomberg School of Public Health, Baltimore, MD, US
147. Institute of Genetic Medicine, Johns Hopkins University School of Medicine, Baltimore, MD, US
148. NORMENT, KG Jebsen Centre for Psychosis Research, Division of Mental Health and Addiction, Institute of Clinical Medicine and Diakonhjemmet Hospital, University of Oslo, Oslo, NO
149. National Institute of Mental Health, Klecany, CZ
150. Discipline of Psychiatry, University of Adelaide, Adelaide, SA, AU
151. Department of Psychiatry and Addiction Medicine, Assistance Publique - Hôpitaux de Paris, Paris, FR
152. Paris Bipolar and TRD Expert Centres, FondaMental Foundation, Paris, FR
153. UMR-S1144 Team 1: Biomarkers of relapse and therapeutic response in addiction and mood disorders, INSERM, Paris, FR
154. Psychiatry, Université Paris Diderot, Paris, FR
155. Psychiatry, University of Pennsylvania, Philadelphia, PA, US
156. Department of Psychiatry, University of Münster, Münster, DE
157. Division of Endocrinology, Children's Hospital Boston, Boston, MA, US
158. Centre for Affective Disorders, Institute of Psychiatry, Psychology and Neuroscience, London, GB
159. Department of Psychiatry & Psychology, Mayo Clinic, Rochester, MN, US
160. School of Medical Sciences, University of New South Wales, Sydney, NSW, AU
161. Department of Human Genetics, University of Chicago, Chicago, IL, US
162. Biometric Psychiatric Genetics Research Unit, Alexandru Obregia Clinical Psychiatric Hospital, Bucharest, RO
163. Institute of Neuroscience and Physiology, University of Gothenburg, Gothenburg, SE
164. INSERM, Paris, FR
165. Department of Medical & Molecular Genetics, King's College London, London, GB
166. Neuroscience Therapeutic Area, Janssen Research and Development, LLC, Titusville, NJ, US
167. Cancer Epidemiology and Prevention, M. Sklodowska-Curie Cancer Center and Institute of Oncology, Warsaw, PL
168. School of Psychology, The University of Queensland, Brisbane, QLD, AU
169. Research Institute, Lindner Center of HOPE, Mason, OH, US
170. Centre for Cognitive Ageing and Cognitive Epidemiology, University of Edinburgh, Edinburgh, GB
171. Human Genetics Branch, Intramural Research Program, National Institute of Mental Health, Bethesda, MD, US
172. Division of Mental Health and Addiction, Oslo University Hospital, Oslo, NO
173. Division of Mental Health and Addiction, University of Oslo, Institute of Clinical Medicine, Oslo, NO
174. Institute of Molecular and Cell Biology, University of Tartu, Tartu, EE
175. Mental Health, Faculty of Medicine and Health Sciences, Norwegian University of Science and Technology - NTNU, Trondheim, NO
176. Psychiatry, St Olavs University Hospital, Trondheim, NO
177. Psychosis Research Unit, Aarhus University Hospital, Risskov, DK
178. Munich Cluster for Systems Neurology (SyNergy), Munich, DE
179. University of Liverpool, Liverpool, GB
180. Psychiatry and Human Genetics, University of Pittsburgh, Pittsburgh, PA, US
181. Mental Health Services in the Capital Region of Denmark, Mental Health Center Copenhagen, University of Copenhagen, Copenhagen, DK
182. Division of Psychiatry, Haukeland Universitetssjukehus, Bergen, NO
183. Faculty of Medicine and Dentistry, University of Bergen, Bergen, NO
184. Human Genetics and Computational Biomedicine, Pfizer Global Research and Development, Groton, CT, US
185. College of Medicine Institute for Genomic Health, SUNY Downstate Medical Center College of Medicine, Brooklyn, NY, US
186. Department of Clinical Genetics, Amsterdam Neuroscience, Vrije Universiteit Medical Center, Amsterdam, NL
187. Department of Neurology and Neurosurgery, McGill University, Faculty of Medicine, Montreal, QC, CA
188. Montreal Neurological Institute and Hospital, Montreal, QC, CA
189. Department of Biomedical and NeuroMotor Sciences, University of Bologna, Bologna, IT
190. Department of Psychiatry, Massachusetts General Hospital, Boston, MA, US
191. Psychiatric and Neurodevelopmental Genetics Unit (PNGU), Massachusetts General Hospital, Boston, MA, US
192. Faculty of Medicine, University of Iceland, Reykjavik, IS
193. Department of Psychiatry, Hospital Namsos, Namsos, NO
194. Department of Neuroscience, Norges Teknisk Naturvitenskapelige Universitet Fakultet for naturvitenskap og teknologi, Trondheim, NO
195. Department of Genetics, University of North Carolina at Chapel Hill, Chapel Hill, NC, US
196. Department of Psychiatry, University of North Carolina at Chapel Hill, Chapel Hill, NC, US
197. Department of Psychiatry, McGill University, Montreal, QC, CA
198. Dept of Psychiatry, Sankt Olavs Hospital Universitetssykehuset i Trondheim, Trondheim, NO
199. Clinical Institute of Neuroscience, Hospital Clinic, University of Barcelona, IDIBAPS, CIBERSAM, Barcelona, ES
200. Institute of Biological Psychiatry, MHC Sct. Hans, Mental Health Services Copenhagen, Roskilde, DK
201. Department of Clinical Medicine, University of Copenhagen, Copenhagen, DK
202. Psychiatry, Indiana University School of Medicine, Indianapolis, IN, US
203. Biochemistry and Molecular Biology, Indiana University School of Medicine, Indianapolis, IN, US

### Authors of the Colombia-US Cross Disorder Collaboration in Psychiatric Genetics Consortium

Susan K. Service 1 MSc,

Cristian Vargas Upegui 2 MD,

Mauricio Castaño Ramírez 3 MD,

Luis Guillermo Agudelo Arango 2 MD,

Ana M. Díaz-Zuluaga 1 MD,

Juanita Melo Espejo 2 MD,

Juan David Palacio 2 MD,

Sergio Ruiz Sánchez 2 BS,

Johanna Valencia 2 BS,

Terri M. Teshiba 1 BA,

Benjamin B. Brodey 4 MD,

Loes Olde Loohuis 1 PhD,

Ruben C. Gur 5 PhD,

Chiara Sabatti 6 PhD,

Javier I. Escobar 7 MD,

Victor I. Reus 8 MD,

Carrie E. Bearden 1 PhD,

Carlos Lopez Jaramillo 2 MD,

Nelson B. Freimer 1 MD.

1. Center for Neurobehavioral Genetics, Semel Institute for Neuroscience and Human Behavior, University of California Los Angeles, Los Angeles, USA

2. Department of Psychiatry, University of Antioquía, Medellín, Colombia

3. Department of Mental Health and Human Behavior, University of Caldas, Manizales, Colombia

4. TeleSage, Inc., Chapel Hill, USA

5. Department of Psychiatry, University of Pennsylvania School of Medicine; Philadelphia, USA

6. Departments of Biomedical Data Science and Statistics, Stanford University, Stanford, USA

7. Department of Psychiatry, Rutgers Robert Wood Johnson Medical School, New Brunswick, USA

8. Department of Psychiatry, University of California San Francisco, San Francisco, USA

### Authors of the International Consortium on Lithium Genetics

| 1. 1 | Bernhard T. | Baune, | Discipline of Psychiatry, | University of Adelaide, Adelaide, | Australia |
| --- | --- | --- | --- | --- | --- |
| 2. 2 | Jan | Fullerton, | Mental Illness (Schofield Group), | Neuroscience Research Australia, Sydney, | Australia |
| 3. | Philip B. | Mitchell, | School of Psychiatry, | University of New South Wales, and Black Dog Institute, Sydney, | Australia |
| 4. | Peter R. | Schofield, | Mental Illness (Schofield Group), | Neuroscience Research Australia, Sydney, | Australia |
| 5. | Naomi R. | Wray, | The University of Queensland, Queensland Brain Institute, | Brisbane, Queensland, | Australia |
| 6. | Adam | Wright, | School of Psychiatry, | University of New South Wales, and Black Dog Institute, Sydney, | Australia |
| 7. | Susanne A. | Bengesser, | Special outpatient center for bipolar affective disorder, | Medical University of Graz, Graz, | Austria |
| 8. | Eva | Reininghaus, | Special outpatient center for bipolar affective disorder, | Medical University of Graz, Graz, | Austria |
| 9. | Claudio E. M. | Banzato, | Department of Psychiatry, | University of Campinas (Unicamp), Campinas, | Brazil |
| 10. | Clarissa | Dantas, | Department of Psychiatry, | University of Campinas (Unicamp), Campinas, | Brazil |
| 11. | Martin | Alda, | Department of Psychiatry, | Dalhousie University, Halifax, Nova Scotia, | Canada |
| 12. | Cristiana | Cruceanu, | Douglas Mental Health University Institute, | McGill University, Montreal, | Canada |
| 13. | Julie | Garnham, | Department of Psychiatry, | Dalhousie University, Halifax, Nova Scotia, | Canada |
| 14. | Paul | Grof, | Mood Disorders Center of Ottawa, |  | Canada |
| 15. | Glenda | MacQueen, | Department of Psychiatry, | University of Calgary, Calgary, | Canada |
| 16. | Guy | Rouleau, | Montreal Neurological Institute and Hospital, | McGill University, Montreal, | Canada |
| 17. | Claire | Slaney, | Department of Psychiatry, | Dalhousie University, Halifax, Nova Scotia, | Canada |
| 18. | Gustavo | Turecki, | Douglas Mental Health University Institute, | McGill University, Montreal, | Canada |
| 19. | L. Trevor | Young, | Department of Psychiatry, | University of British Columbia, Vancouver, | Canada |
| 20. | Carlos A. | López Jaramillo, | Department of Psychiatry, | University of Antioquia, Medellín, Medellín, | Colombia |
| 21. | Tomás | Novák, | Prague Psychiatric Center and 3rd Faculty of Medicine, | Charles University, Prague, | Czech Republic |
| 22. | Pavla | Stopkova, | Prague Psychiatric Center and 3rd Faculty of Medicine, | Charles University, Prague, | Czech Republic |
| 23. | Frank | Bellivier, | INSERM UMR-S 1144 - Pôle de Psychiatrie, | AP-HP, Groupe Hospitalier Lariboisière-F. Widal, Paris, | France |
| 24. | Clara | Brichant-Petitjean, | INSERM UMR-S 1144 - Pôle de Psychiatrie, | AP-HP, Groupe Hospitalier Lariboisière-F. Widal, Paris, | France |
| 25. | Bruno | Etain, | Inserm U955, Psychiatrie Génétique, | Créteil, | France |
| 26. | Bruno | Etain, | Université Paris Est, Faculté de Médecine, | Créteil, | France |
| 27. | Bruno | Etain, | Fondation FondaMental, | Créteil, | France |
| 28. | Bruno | Etain, | Assistance Publique-Hôpitaux de Paris, | Hôpital Albert Chenevier - Henri Mondor, Pôle de Psychiatrie, Créteil, | France |
| 29. | Sébastien | Gard, | Service de psychiatrie, | Hôpital Charles Perrens, Bordeaux, | France |
| 30. | Stéphane | Jamain, | Inserm U955, Psychiatrie Génétique, | Créteil, | France |
| 31. | Stéphane | Jamain, | Université Paris Est, Faculté de Médecine, | Créteil, | France |
| 32. | Stéphane | Jamain, | Fondation FondaMental, | Créteil, | France |
| 33. | Jean-Pierre | Kahn, | Service de Psychiatrie et Psychologie Clinique, | Centre Hospitalier Universitaire de Nancy, Nancy, | France |
| 34. | Jean-Pierre | Kahn, | Université de Lorraine, | Nancy, | France |
| 35. | Marion | Leboyer, | Inserm U955, Psychiatrie Génétique, | Créteil, | France |
| 36. | Marion | Leboyer, | Université Paris Est, Faculté de Médecine, | Créteil, | France |
| 37. | Marion | Leboyer, | Fondation FondaMental, | Créteil, | France |
| 38. | Marion | Leboyer, | Assistance Publique-Hôpitaux de Paris, | Hôpital Albert Chenevier - Henri Mondor, Pôle de Psychiatrie, Créteil, | France |
| 39. | Mazda | Adli, | Department of Psychiatry and Psychotherapy, Charité - Universitätsmedizin Berlin, | Campus Charité Mitte & Fliedner Klinik Berlin, | Germany |
| 40. | Mazda | Adli, | Fliedner Klinik Berlin, | Berlin, | Germany |
| 41. | Michael | Bauer, | Department of Psychiatry and Psychotherapy, | University Hospital Carl Gustav Carus, Technische Universität Dresden, Dresden, | Germany |
| 42. | Sven | Cichon, | Institute of Human Genetics, Department of Genomics, Life and Brain Center, | University of Bonn, Bonn, | Germany |
| 43. | Sven | Cichon, | Institute of Neuroscience and Medicine (INM-1), Genomic Imaging, | Research Center Juelich, Juelich, | Germany |
| 44. | Franziska | Degenhardt, | Institute of Human Genetics, Department of Genomics, Life and Brain Center, | University of Bonn, Bonn, | Germany |
| 45. | Peter | Falkai, | Department of Psychiatry and Psychotherapy, | Ludwig-Maximilians-University Munich, Munich, | Germany |
| 46. | Oliver | Gruber, | Department of Psychiatry and Psychotherapy, | Georg-August University Göttingen, Göttingen, | Germany |
| 47. | Urs | Heilbronner, | Department of Psychiatry and Psychotherapy, | Georg-August University Göttingen, Göttingen, | Germany |
| 48. | Per | Hoffmann, | Institute of Human Genetics, Department of Genomics, Life and Brain Center, | University of Bonn, Bonn, | Germany |
| 49. | Per | Hoffmann, | Institute of Neuroscience and Medicine (INM-1), Genomic Imaging, | Research Center Juelich, Juelich, | Germany |
| 50. | Sarah | Kittel-Schneider, | Department of Psychiatry, Psychosomatics, and Psychotherapy, | University of Würzburg, Würzburg, | Germany |
| 51. | Markus | Nöthen, | Institute of Human Genetics, Department of Genomics, Life and Brain Center, | University of Bonn, Bonn, | Germany |
| 52. | Andrea | Pfennig, | Department of Psychiatry and Psychotherapy, | University Hospital Carl Gustav Carus, Technische Universität Dresden, Dresden, | Germany |
| 53. | Daniela | Reich-Erkelenz, | Department of Psychiatry and Psychotherapy, | Ludwig-Maximilians-University Munich, Munich, | Germany |
| 54. | Andreas | Reif, | Department of Psychiatry, Psychosomatics, and Psychotherapy, | University of Würzburg, Würzburg, | Germany |
| 55. | Marcella | Rietschel, | Department of Genetic Epidemiology in Psychiatry, | Central Institute of Mental Health, Mannheim, | Germany |
| 56. | Thomas G. | Schulze, | Department of Psychiatry and Psychotherapy, | Georg-August University Göttingen, Göttingen, | Germany |
| 57. | Florian | Seemüller, | Department of Psychiatry and Psychotherapy, | Ludwig-Maximilians-University Munich, Munich, | Germany |
| 58. | Thomas | Stamm, | Department of Psychiatry and Psychotherapy, | Charité - Universitätsmedizin Berlin, Campus Charité Mitte, Berlin, | Germany |
| 59. | Raffaella | Ardau, | Unit of Clinical Pharmacology, Hospital University Agency, | University of Cagliari, Cagliari, | Italy |
| 60. | Caterina | Chillotti, | Unit of Clinical Pharmacology, Hospital University Agency, | University of Cagliari, Cagliari, | Italy |
| 61. | Maria | Del Zompo, | Department of Biomedical Sciences, | University of Cagliari, Cagliari, | Italy |
| 62. | Maria | Del Zompo, | Unit of Clinical Pharmacology, Hospital University Agency, | University of Cagliari, Cagliari, | Italy |
| 63. | Mario | Maj, | Department of Psychiatry, | University of Naples, SUN, Naples, | Italy |
| 64. | Mirko | Manchia, | Department of Biomedical Sciences, | University of Cagliari, Cagliari, | Italy |
| 65. | Palmiero | Monteleone, | Department of Psychiatry, | University of Naples, SUN, Naples, | Italy |
| 66. | Giovanni | Severino, | Department of Biomedical Sciences, | University of Cagliari, Cagliari, | Italy |
| 67. | Alessio | Squassina, | Department of Biomedical Sciences, | University of Cagliari, Cagliari, | Italy |
| 68. | Alfonso | Tortorella, | Department of Psychiatry, | University of Naples, SUN, Naples, | Italy |
| 69. | Kazufumi | Akiyama, | Department of Biological Psychiatry and Neuroscience, | Dokkyo Medical University School of Medicine, Mibu, | Japan |
| 70. | Kazufumi | Akiyama, | The Japanese Collaborative Group on the Genetics of Lithium Response in Bipolar Disorder, |  | Japan |
| 71. | Ryota | Hashimoto, | Molecular Research Center for Children’s Mental Development, United Graduate School of Child Development, Osaka University, | Osaka University, Osaka, | Japan |
| 72. | Ryota | Hashimoto, | The Japanese Collaborative Group on the Genetics of Lithium Response in Bipolar Disorder, |  | Japan |
| 73. | ~~Nakao~~ | ~~Iwata,~~ | ~~Department of Psychiatry,~~ | ~~Fujita Health University School of Medicine, Toyoake,~~ | ~~Japan~~ |
| 74. | Tadafumi | Kato, | The Japanese Collaborative Group on the Genetics of Lithium Response in Bipolar Disorder, |  | Japan |
| 75. | Tadafumi | Kato, | Laboratory for Molecular Dynamics of Mental Disorders, | RIKEN Brain Science Institute, Saitama, | Japan |
| 76. | Ichiro | Kusumi, | The Japanese Collaborative Group on the Genetics of Lithium Response in Bipolar Disorder, |  | Japan |
| 77. | Ichiro | Kusumi, | Department of Psychiatry, | Hokkaido University Graduate School of Medicine, Sapporo, | Japan |
| 78. | Takuya | Masui, | The Japanese Collaborative Group on the Genetics of Lithium Response in Bipolar Disorder, |  | Japan |
| 79. | Takuya | Masui, | Department of Psychiatry, | Hokkaido University Graduate School of Medicine, Sapporo, | Japan |
| 80. | Norio | Ozaki, | The Japanese Collaborative Group on the Genetics of Lithium Response in Bipolar Disorder, |  | Japan |
| 81. | Norio | Ozaki, | Department of Psychiatry, | Nagoya University Graduate School of Medicine, Nagoya, | Japan |
| 82. | Piotr | Czerski, | Psychiatric Genetic Unit, | Poznan University of Medical Sciences, Poznan, | Poland |
| 83. | Joanna | Hauser, | Psychiatric Genetic Unit, | Poznan University of Medical Sciences, Poznan, | Poland |
| 84. | Sebastian | Kliwicki, | Department of Adult Psychiatry, | Poznan University of Medical Sciences, Poznan, | Poland |
| 85. | Janusz K. | Rybakowski, | Department of Adult Psychiatry, | Poznan University of Medical Sciences, Poznan, | Poland |
| 86. | Maria | Grigoroiu-Serbanescu, | Biometric Psychiatric Genetics Research Unit, | Alexandru Obregia Psychiatric Hospital, Bucharest, | Romania |
| 87. | Bárbara | Arias, | Department of Biologia Animal, Unitat d'Antropologia, Facultat de Biologia, Universitat de Barcelona, IBUB, CIBERSAM, | Instituto de Salud Carlos III, Barcelona, Catalonia, | Spain |
| 88. | Antonio | Benabarre, | Bipolar Disorders Program, Institute of Neuroscience, Hospital Clinic, | University of Barcelona, IDIBAPS, CIBERSAM, Barcelona, Catalonia, | Spain |
| 89. | Francesc | Colom, | Bipolar Disorders Program, Institute of Neuroscience, Hospital Clinic, | University of Barcelona, IDIBAPS, CIBERSAM, Barcelona, Catalonia, | Spain |
| 90. | Esther | Jiménez, | Bipolar Disorders Program, Institute of Neuroscience, Hospital Clinic, | University of Barcelona, IDIBAPS, CIBERSAM, Barcelona, Catalonia, | Spain |
| 91. | Marina | Mitjans, | Department of Biologia Animal, Unitat d'Antropologia, Facultat de Biologia, Universitat de Barcelona, IBUB, CIBERSAM, | Instituto de Salud Carlos III, Barcelona, Catalonia, | Spain |
| 92. | Eduard | Vieta, | Bipolar Disorders Program, Institute of Neuroscience, Hospital Clinic, | University of Barcelona, IDIBAPS, CIBERSAM, Barcelona, Catalonia, | Spain |
| 93. | Lena | Backlund, | Department of Molecular Medicine and Surgery, | Karolinska Institutet and Center for Molecular Medicine, Karolinska University Hospital, Stockholm, | Sweden |
| 94. | Lena | Backlund, | Department of Clinical Neuroscience, Centre for Psychiatric Research and Education, | Karolinska Institutet, The Clinic for Affective Disorders, Karolinska University Hospital, Stockholm, | Sweden |
| 95. | Louise | Frisén, | Department of Molecular Medicine and Surgery, | Karolinska Institutet and Center for Molecular Medicine, Karolinska University Hospital, Stockholm, | Sweden |
| 96. | Louise | Frisén, | Department of Clinical Neuroscience, Centre for Psychiatric Research and Education, | Karolinska Institutet, The Clinic for Affective Disorders, Karolinska University Hospital, Stockholm, | Sweden |
| 97. | Catharina | Lavebratt, | Department of Molecular Medicine and Surgery, | Karolinska Institutet and Center for Molecular Medicine, Karolinska University Hospital, Stockholm, | Sweden |
| 98. | Lina | Martinsson, | Department of Molecular Medicine and Surgery, | Karolinska Institutet and Center for Molecular Medicine, Karolinska University Hospital, Stockholm, | Sweden |
| 99. | Lina | Martinsson, | Department of Clinical Neuroscience, Centre for Psychiatric Research and Education, | Karolinska Institutet, The Clinic for Affective Disorders, Karolinska University Hospital, Stockholm, | Sweden |
| 100. | Urban | Ösby, | Department of Molecular Medicine and Surgery, | Karolinska Institutet and Center for Molecular Medicine, Karolinska University Hospital, Stockholm, | Sweden |
| 101. | Martin | Schalling, | Department of Molecular Medicine and Surgery, | Karolinska Institutet and Center for Molecular Medicine, Karolinska University Hospital, Stockholm, | Sweden |
| 102. | Jean-Michel | Aubry, | Départment de Psychiatrie, | HUG - Hôpitaux Universitaires de Genève, Geneva, | Switzerland |
| 103. | Sven | Cichon, | Division of Medical Genetics, | Department of Biomedicine, University of Basel, Basel, | Switzerland |
| 104. | Alexandre | Dayer, | Départment de Psychiatrie, | HUG - Hôpitaux Universitaires de Genève, Geneva, | Switzerland |
| 105. | Alexandre | Dayer, | Department of Basic Neurosciences, | University of Geneva Medical School, Geneva, | Switzerland |
| 106. | Per | Hoffmann, | Division of Medical Genetics, | Department of Biomedicine, University of Basel, Basel, | Switzerland |
| 107. | Audrey | Nallet, | Department of Mental Health and Psychiatry, | Hôpitaux Universitaires de Genéve, Geneva, | Switzerland |
| 108. | Hsi-Chung | Chen, | Department of Psychiatry & Center of Sleep Disorders, | National Taiwan University Hospital, Taipei, | Taiwan |
| 109. | Po-Hsiu | Kuo, | Institute of Epidemiology and Preventive Medicine, | National Taiwan University, Taipei, | Taiwan |
| 110. | David | Cousins, | Campus for Ageing and Vitality, | Newcastle University, Newcastle, | United Kingdom |
| 111. | Nirmala | Akula, | Human Genetics Branch, | National Institute of Mental Health and Human Services, Bethesda, MD, | United States |
| 112. | Joanna M. | Biernacka, | Department of Psychiatry and Psychology, | Mayo Clinic, Rochester, MN, | United States |
| 113. | Joanna M. | Biernacka, | Department of Health Sciences Research, | Mayo Clinic, Rochester, MN, | United States |
| 114. | Elise T. | Bui, | Human Genetics Branch, | National Institute of Mental Health and Human Services, Bethesda, MD, | United States |
| 115. | J. Ray | DePaulo, | Department of Psychiatry and Behavioral Sciences, | Johns Hopkins University, Baltimore, MD, | United States |
| 116. | Sevilla D. | Detera-Wadleigh, | Human Genetics Branch, | National Institute of Mental Health and Human Services, Bethesda, MD, | United States |
| 117. | Mark A. | Frye, | Department of Psychiatry and Psychology, | Mayo Clinic, Rochester, MN, | United States |
| 118. | Fernando S. | Goes, | Department of Psychiatry and Behavioral Sciences, | Johns Hopkins University, Baltimore, MD, | United States |
| 119. | Rebecca | Hoban, | Department of Psychiatry, | University of California San Diego, | United States |
| 120. | Rebecca | Hoban, | Department of Psychiatry, | VA San Diego Healthcare System, | United States |
| 121. | Liping | Hou, | Human Genetics Branch, | National Institute of Mental Health and Human Services, Bethesda, MD, | United States |
| 122. | Layla | Kassem, | Human Genetics Branch, | National Institute of Mental Health and Human Services, Bethesda, MD, | United States |
| 123. | John R. | Kelsoe, | Department of Psychiatry, | University of California San Diego, San Diego, CA, | United States |
| 124. | John R. | Kelsoe, | Department of Psychiatry, | VA San Diego Healthcare System, San Diego, CA, | United States |
| 125. | Gonzalo | Laje, | Human Genetics Branch, | National Institute of Mental Health and Human Services, Bethesda, MD, | United States |
| 126. | Gonzalo | Laje, | Washington Behavioral Medicine Associates, LLC, | Chevy Chase, MD, | United States |
| 127. | Gonzalo | Laje, | Maryland Institute for Neuroscience & Development (MIND), | Chevy Chase, MD, | United States |
| 128. | Susan G. | Leckband, | Department of Psychiatry, | University of California San Diego, San Diego, CA, | United States |
| 129. | Susan G. | Leckband, | Department of Pharmacy, | VA San Diego Healthcare System, San Diego, CA, | United States |
| 130. | Susan G. | Leckband, | Skaggs School of Pharmacy and Pharmaceutical Sciences, | University of California, San Diego, CA, | United States |
| 131. | Michael J. | McCarthy, | Department of Psychiatry, | VA San Diego Healthcare System, San Diego, CA, | United States |
| 132. | Francis J. | McMahon, | Human Genetics Branch, | National Institute of Mental Health and Human Services, Bethesda, MD, | United States |
| 133. | Francis | Mondimore, | Department of Psychiatry and Behavioral Sciences, | Johns Hopkins University, Baltimore, MD, | United States |
| 134. | Roy H. | Perlis, | Department of Psychiatry, | Massachusetts General Hospital and Harvard Medical School, Boston, MA, | United States |
| 135. | James B. | Potash, | Department of Psychiatry, | University of Iowa, Iowa City, IA, | United States |
| 136. | Thomas G. | Schulze, | Department of Psychiatry and Behavioral Sciences, | Johns Hopkins University, Baltimore, MD, | United States |
| 137. | Thomas G. | Schulze, | Human Genetics Branch, | National Institute of Mental Health and Human Services, Bethesda, MD, | United States |
| 138. | Barbara | Schweizer, | Department of Psychiatry and Behavioral Sciences, | Johns Hopkins University, Baltimore, MD, | United States |
| 139. | Lisa R. | Seymour, | Department of Psychiatry, | Mayo Clinic, Rochester, MN, | United States |
| 140. | Jordan W. | Smoller, | Department of Psychiatry, | Massachusetts General Hospital and Harvard Medical School, Boston, MA, | United States |
| 141. | Jo | Steele, | Human Genetics Branch, | National Institute of Mental Health and Human Services, Bethesda, MD, | United States |
| 142. | Sarah | Tighe, | Department of Psychiatry, | University of Iowa, Iowa City, IA, | United States |
| 143. | Peter P. | Zandi, | Department of Mental Health, | Johns Hopkins Bloomberg School of Public Health, Baltimore, MD, | United States |
